# Phenotypic and genetic sex differences in LDL-C and statin treatment in All of Us

**DOI:** 10.64898/2026.01.26.26344867

**Authors:** Jaclyn L. Liquori, Xiaohe Tian, Shaila A. Musharoff

## Abstract

Low-density lipoprotein cholesterol (LDL-C) is a well-established and modifiable risk factor for cardiovascular diseases (CVDs), which have been the leading cause of mortality among individuals in the United States since 1921. Despite known sex differences in CVD presentation and outcomes, women remain underdiagnosed and undertreated relative to men. Using genotype and phenotype data from the All of Us (AoU) Research Program, we assessed sex-specific differences in LDL-C based on measured levels, statin treatment rates and efficacy, age-related patterns, and genetic factors as reflected by GWAS, XWAS, GxSex interaction, and heritability. We find that the median LDL-C measurements of women are consistently higher than those of men across most age strata, regardless of statin treatment, and that this difference is greatest after 40 years of age. In addition, women are treated with statins less often than their male counterparts and appear to respond less effectively to statins. This is particularly true for Black women, who had the highest statin-treated median LDL-C of any race-sex stratum. Our sex-stratified GWAS results recapitulate known sex-differentiated effects at the *APOE/APOC1* locus, and our male-stratified XWAS identified an LDL-C associated SNP in the *PTCHD1-AS* region. While heritability and additive genetic variability of LDL-C do not demonstrate sex differences, residual and phenotypic variability do. This suggests that environmental factors may drive variation in LDL-C levels and statin treatment rates across sex and self-identified race groups. These findings highlight the need for sex- and population-specific strategies in genetic studies and LDL-C treatment.

## Introduction

CVDs are the leading cause of death of both men and women worldwide [1]. Despite this, CVD remains underdiagnosed and undertreated in women [2]; when women are diagnosed and treated, they experience worse outcomes relative to men [3]. CVD risk varies by race/ethnicity, with Black and Asian individuals having a higher risk of developing CVD relative to White individuals [4]. Various blood lipids, including LDL-C, for which population-mean levels vary by genetic ancestry [5], [6], contribute to the risk of developing CVD. LDL-C is well-known for its role in the development of atherosclerosis, a condition that contributes to the development of CVDs.

Moreover, known disparities in LDL-C management exist between sexes. First, women receive routine blood lipid screening later in life than men; the current clinical recommendation is to begin routine blood lipid screening for women at 45 years old and men at 35 years old [7]. This timing is particularly consequential for women as those aged 40–60 are experiencing peri-menopause and menopause, a period marked by lipid changes and rising LDL-C levels; the later screening age therefore delays when women may be offered LDL-C modification options. Second, women report higher rates of adverse effects related to statin use than men do, which could lead to the refusal or discontinuation of statin use if they are prescribed [8].

Research has established that the presentation and progression of CVD differs between women and men [2], [9], and previous studies have demonstrated the utility of sex-stratified genomic analysis of sexually differentiated diseases [10], [11]. Studies have examined the sexually dimorphic expression of various risk factors for CVD, including adipose tissue distribution, blood pressure, and multiple biomarkers important to cardiac health [12], [13], [14]. While the influence of potential sex-specific differences in genetic risk factors has been established [15], comprehensive sex-stratified analyses in ancestrally diverse populations remain limited. Existing studies have been restricted mainly to the White British UK Biobank cohort [16] and are often limited by the under-recruitment of female participants [17] or small sample sizes [18]. Large-scale LDL-C GWAS efforts [19] and sex-stratified meta-analyses [20] have identified hundreds of lipid-associated loci and begun to characterize sex-biased genetic effects at scale. However, these studies differ in phenotype and scope. Furthermore, these meta-analyses do not often examine sex differences in LDL-C phenotype, statin treatment rates, or treatment efficacy. While some recent work has examined lipid genetics in more diverse cohorts [21], [22], [23], [5], and recent work has begun to address sex-specific lipid genetics in the AoU Research Program [24], comprehensive analysis focusing specifically on LDL-C remains incomplete, particularly with respect to examining treatment responses across diverse racial groups. While studies focusing exclusively on European or self-identified White individuals are often performed for increased statistical power and consistency of results, these results are not always applicable to other population groups [25], [26], and applying these results to different groups may lead to even more disparity in health outcomes [27]. GWAS are used to identify genetic variation, such as single nucleotide polymorphisms (SNPs), associated with traits. The sex-stratification of GWAS has the potential to elucidate differences in trait-associated genetic variants between women and men [28], [29], [30], [31], [32].

Furthermore, the X chromosome (chrX), despite making up approximately 5% of the human genome [33], is frequently excluded from GWAS pipelines due to technical challenges presented by its unique inheritance pattern [34]; therefore, its role in complex traits in general—and in lipid regulation specifically— remains under-studied. X chromosome-wide association studies (XWAS) have been able to identify associated loci for multiple complex traits [35], [36], [37], including cardiometabolically relevant traits such as heart failure [38], fasting insulin [39], and lipids [40]. However, XWAS analyses of lipid-related traits are commonly performed in sex-pooled groups [41] which could miss potential sex-specific associations [42], [43]. Additionally, sex and statin treatment status may influence the genetic architecture of LDL-C, including at lipid-associated loci [44], including on the X chromosome where genetic effects between females and males may differ. Accounting for these factors may therefore improve the detection and interpretation of genetic associations with LDL-C and how they may be sex-differentiated. More recently, meta-analyses have begun to investigate chrX in sex-stratified lipid analyses that include statin-treated individuals at biobank scale [20]. However, the availability of X-chromosome summary statistics remains limited [44]. This limited availability of XWAS summary statistics can constrain the incorporation of chrX into large-scale meta-analyses. To our knowledge, no study has performed a sex-stratified XWAS of statin-adjusted LDL-C in AoU.

This study investigates whether there are sex-specific genetic variants that influence LDL-C levels, as well as sex differences in the LDL-C phenotype itself. We use genotype and phenotype data from the v7 release of the AoU Research Program to examine if LDL-C levels, statin efficacy, age-related changes in LDL-C levels, genetic associations and their effect sizes, and SNP-based heritability differ between women and men. Specifically, we characterize sex differences in the distribution and central tendency of LDL-C by statin-treatment status, and we estimate SNP-based heritability and its underlying variance components separately in women and men to assess whether additive genetic architecture of LDL-C differs by sex. We focus specifically on LDL-C given its clinical importance as a modifiable cardiovascular risk factor and extend previous genomic analyses by examining biobank-level treatment patterns. Due to their majority representation in the AoU dataset, we focus on cisgender women and cisgender men, who we refer to as “women” or “female” and “men” or “male”, respectively, using reported sex at birth as the analysis variable and the terms women and men as humanizing language.

Importantly, conducting these analyses within a single cohort provides a complementary contribution to previous multi-cohort LDL-C studies. A single-cohort design allows phenotype definition, statin treatment ascertainment, covariate adjustment, and autosomal and X-chromosome analyses to be performed consistently across all participants. Furthermore, we designed this study to increase diversity in research analyses by including individuals of diverse self-reported racial and ethnic identities. By assessing sex differences across phenotypic, treatment, and genomic domains within a racially diverse cohort of individuals, we may better understand the multifactor contributors to LDL-C variation and CVD risk.

## Methods

### Study group

#### Combined analysis group filtration

Using the AoU dataset, we restricted analysis to the largest groups of individuals based on sex and gender: cisgender women and cisgender men, defined as individuals who identify as the gender which is most commonly associated with their self-reported assigned sex at birth, who we refer to as “women” or “female” and “men” or “male”, respectively. These individuals’ self-reported racial and ethnic identities include Asian, Black or African American, Hispanic or Latino, Middle Eastern or North African, Native Hawaiian or Other Pacific Islander, White, “More than one population”, and “None of these”.

These individuals had whole genome sequencing (WGS) information available as well as Electronic Health Record (EHR) data containing LDL-C measurements, age at the time of measurement, and statin prescription data. We restricted the cohort of interest to individuals between the ages of 21 and 80 at the time of measurement, as the sample size of individuals outside this range was not large enough to support well-powered subgroup analyses. Finally, we searched the EHR data of each individual for a diagnosis of familial hypercholesterolemia (OMOP concept ID 4134862). This condition can lead to extremely elevated LDL-C in very young people. We removed individuals with a diagnosis of familial hypercholesterolemia from the analysis group. After the above filtration steps, the study population (N = 86,700) was comprised of cisgender women (n = 54,189) and cisgender men (n = 32,511) of diverse self-reported racial and ethnic identities, and we refer to this group as the combined analysis group.

#### Race and ethnicity variables

We restricted race-stratified analysis to the two largest groups based on self-reported racial identities, “White” and “Black or African American”. We stratified the combined analysis group by sex and self-reported racial identities. We do not use race and ethnicity as a proxy for genetic ancestry in this study, following guidelines from The National Academies of Sciences, Engineering, and Medicine [45]. Instead, self-identified race is used only in phenotypic analyses which aim to capture social and structural factors—including prescribing patterns and systemic barriers—rather than biological differences.

### LDL-C phenotype definition and statistical comparisons

#### Phenotype refinement

We defined the LDL-C phenotype utilizing an existing pipeline [46], which was originally developed to demonstrate cross-cohort analysis of AoU and UK Biobank data and is available as a featured workspace on the AoU Researcher Workbench. This pipeline has since been used in subsequent AoU genomic studies [22], [47]. This phenotype is built as follows. We grouped Serum or Plasma LDL-C measurement methods based on LOINC codes 13457-7, 2089-1, 18262-6, 18261-8, and 49132-4 (measured by calculation, unspecified method, direct assay, ultracentrifugation, and electrophoresis, respectively), ensured that measurement units are consistent (mg/dL), limited LDL-C measurements to the most recent measurement, and ensured that the statin usage indicator is accurate for time of measurement in the following way. Statin use was determined using EHR drug exposure data. An individual was classified as receiving statin therapy if their LDL-C measurement date fell within the interval bounded by the dates of their first and last recorded statin prescriptions. An individual was considered untreated if they had no statin prescription recorded at any time, or if their LDL-C measurement date fell before the first prescription or after the last recorded prescription.

We then restricted LDL-C measurements to a biologically plausible range of 20 mg/dL to 300 mg/dL. We restricted the upper bound of LDL-C to 300 mg/dL to eliminate extreme outliers, such as undiagnosed homozygous FH (often ≥ 400 mg/dL) [48], [49], and severe measurement errors. Though this cutoff does not fully exclude undiagnosed heterozygous FH, it is a supplementary filter for the most extreme tail of the distribution minimizing the influence of extreme outliers.

Further refinement included adjusting LDL-C measurement to account for statin usage, specifically by dividing the LDL-C measurement by 0.7. This standard adjustment estimates the untreated LDL-C level for a statin-treated individual and is based on a conservative estimate of statin efficacy [50]. We use the untransformed phenotype (i.e., without log, rank-based inverse normal, or other transformations) to preserve interpretability of estimated effects and facilitate comparisons with clinical literature.

We additionally examine maximum LDL-C measurements that are refined as above. Maximum LDL-C is, by definition, the most extreme observation, potentially skewing analyses of maximum LDL-C towards biological extremes, non-fasting measurements, and illness states. Therefore, we use the most recent measurement as our primary LDL-C phenotype, consistent with the phenotype definition used in other AoU LDL-C analyses [22], [46], [47], [51].

#### Statistical methods for phenotype comparisons

We examined the distribution of LDL-C measurements for both women and men by estimating the probability density function (PDF) for each group using kernel density estimation (KDE). We then interpolated the resulting densities by using the approx() function in R, and normalized the density values by dividing them by their sum to convert them to probabilities. To quantify differences in the overall distributional shape between sexes (beyond central tendency), we calculated the Kullback-Leibler (KL) divergence between female and male probability density functions. KL divergence measures how one probability distribution diverges from a second, expected probability distribution, with higher values indicating greater dissimilarity in shape.

Given the skew of the LDL-C phenotype, we chose the Mann-Whitney U (MWU) test to assess significant differences between women and men. The MWU test is a non-parametric test that determines significant differences between the distributions of two independent groups and does not assume normality. After pooling samples from both groups, the MWU test produces a test statistic called *U* with a known distribution derived from the ranks assigned to all observations. We calculated the MWU test statistic in R using the wilcox.test() function.

A drawback to non-parametric methods such as the MWU test is that they can be sensitive to ties or instances where two or more observations have the same value. There are many repeated LDL-C measurements and, thus, many ties that can affect the data ranking. To account for the ties in the data and their influence on the MWU statistic, we generated a Hodges-Lehmann (HL) estimate. While the MWU test indicates a significant difference in central tendency, the HL estimator provides a quantified estimate of the difference in the median between the two groups. This estimate is non-parametric and is complementary to a MWU test. The HL estimator also accounts for ties within data by adjusting the ranks such that each observation will receive the average rank of their position. Because the HL estimator is based on pairwise average rank, ties still affect the estimate. Still, they do not invalidate it as the averages reflect the central tendency of the data. We used the HodgesLehmann() function from the ‘DescTools’ R package [52] to obtain the HL estimator by conducting a bootstrap analysis with 1,000 trials sampled from the data. We then averaged the difference in medians across all trials to obtain the HL estimate.

#### Phenotype sensitivity analyses

To test the robustness of the primary phenotype (most recent LDL-C, filtered to 20-300 mg/dL), we repeated the sex-difference median analyses using maximum LDL-C measurements across three filtration ranges: <1000 mg/dL, 10–500 mg/dL, and the primary 20–300 mg/dL range. For each combination of phenotype (most recent versus maximum), filtration range, and subgroup (all individuals, statin-treated, untreated), we computed female and male medians, HL estimates of the median difference with 95% confidence intervals, MWU p-values, and KL divergences to assess distributional shape differences between sexes. These sensitivity results are reported separately for unadjusted and statin-adjusted LDL-C. Because maximum LDL-C is, by definition, greater than or equal to an individual’s most recent LDL-C value, and the same filtration bounds were applied to both phenotypes, the two phenotype definitions do not necessarily retain the same set of individuals. As a result, sample sizes differ slightly between the most recent and maximum LDL-C analyses within each subgroup and filtration range.

#### Sensitivity analysis for LDL-C measurement method

To assess the potential impact of LDL-C measurement method on GWAS results, we compared two linear models using the same GWAS phenotype (statin-adjusted, untransformed LDL-C) and covariates (sex, age, age squared, and the first ten genetic PCs). The full model includes measurement method (categorical: calculation, direct, ultracentrifugation, electrophoresis, and unspecified) while the reduced model omitted it. We calculated the partial R^2^ for method as:

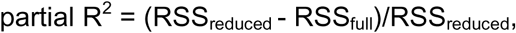

and recorded the change in total model R^2^.

### Genome-wide association studies

#### Genomic data preparation

We utilized the AoU GRCh38/hg38 v7 ACAF call sets for all genomic analyses in our study. AoU built these call sets to contain variants in the genomic data with population-specific allele count (AC) greater than 100 or allele frequency (AF) greater than 1%. Using PLINK1.9 [53], [54], we filtered and pruned SNPs for analysis. We restricted the variants of interest to autosomal biallelic SNPs that we additionally filtered with the following parameters: minor allele frequency (MAF) > 0.05 to restrict to common variants for adequate statistical power, minor allele count (MAC) threshold of > 100 to prevent the inclusion of variants with insufficient data, and Hardy-Weinberg Equilibrium (HWE) > 1e-6 to retain high-quality SNPs. In addition, we performed LD pruning to remove one of a pair of SNPs whose r² value exceeded 0.2, which we performed using a sliding window of 50 SNPs, advancing by 5 SNPs at a time. We obtained predicted genetic ancestry for each individual from the AoU ancestry_preds.tsv file which groups individuals based on their genetic similarity to reference populations from gnomAD, HGDP, and 1000 Genomes. The AoU platform includes genetic principal components (PCs) obtained from performing a principal component analysis (PCA) for the entire non-filtered cohort. These PCs are in the ancestry_preds.tsv file, and included in appropriate analyses.

#### Combined analysis group GWAS

We conducted a GWAS on the entirety of the combined analysis group to compare results to existing GWAS to ensure that our analysis could reproduce known results from the AoU dataset [22] and the TOPMed dataset [23]. We performed the GWAS using REGENIE, a fast and effective linear mixed model analysis program [55]. We conducted the GWAS on the entire combined analysis group (N = 86,700) with the first ten genetic PCs, age, age squared, and sex as covariates and statin-adjusted LDL-C measurements as the phenotype. We calculated the genomic inflation factor (λ) [56], which measures the degree of inflation in the test statistics, where values closer to 1 suggest no significant inflation.

To further ensure the consistency of our GWAS results, we compared effect size estimates at SNPs reaching genome-wide significance in both the pooled GWAS and the TOPMed GWAS. The AoU researchers utilized this quality control step when examining the consistency of their dataset [22], and we repeated it with the combined analysis group to compare the efficacy of the model in our study.

#### Sex-stratified GWAS

We stratified the combined analysis group into women and men only. We then performed a GWAS on both subsets using age, age squared, and the first ten genetic PCs as covariates. After running the GWAS, we applied Bonferroni correction to adjust the p-values of SNPs. We used a genome-wide significance threshold of p < 4.94e-8 (0.05 / 1,011,764 variants tested), where 1,011,764 represents the largest number of variants tested across the three GWAS.

We annotated genome-wide significant SNPs from each analysis using biomaRt [57] to retrieve the nearest protein-coding gene from Ensembl (GRCh38). For each SNP, we first queried a 10kb window around its position; if no protein-coding gene was found, we expanded the window to 500kb and selected the closest gene based on the minimum distance to the transcription start or end site. We then obtained rsIDs by querying the NCBI dbSNP database through its E-utils API using the chromosome and position.

#### Genotype-by-sex interaction analysis

We performed a formal genotype-by-sex (GxSex) interaction GWAS using REGENIE in the combined analysis group. We performed the GWAS with statin-adjusted LDL-C as the phenotype and age, age squared, sex, and the first ten genetic PCs as covariates. We tested for GxSex interactions using the ‘--interaction’ flag in STEP 2 of REGENIE, treating sex as a categorical variable with females as the reference group; therefore, the interaction effect represents the male-minus-female difference in per-allele effect, and we report the SNP-by-sex estimated effect. We determined significance using a Bonferroni-adjusted genome-wide significance threshold of p < 4.94e-8 (0.05 / 1,011,764 variants). We additionally evaluated interaction estimates at the 14 lead loci from the sex-difference analysis using a locus-level threshold as described below.

### X chromosome-wide association study

#### Chromosome X data preparation

Because the X chromosome requires additional quality control not applicable to autosomes to account for hemizygosity in males, we performed a separate quality control (QC) pipeline for chrX prior to XWAS. We identified genetically related individuals using the AoU relatedness_flagged_samples.tsv auxiliary file, which contains the minimum set of individuals removed to obtain an unrelated group, and excluded these individuals from downstream chrX analyses. We obtained self-reported sex at birth and predicted ancestry for each individual based on genetic PCs in the ancestry_preds.tsv file. Using the chrX PLINK files from the AoU v7.1 ACAF threshold call set, we assessed concordance between reported sex and chrX data separately within each ancestry group for sex-concordance QC. For each ancestry group, we excluded the pseudoautosomal regions (PARs), restricted to biallelic SNPs, filtered to MAF > 0.05, and LD-pruned the resulting variants (50 window, 5 step, r^2^ > 0.2) using PLINK 2.0 [58]. We then thinned the LD-pruned set to 20,000 variants and used these variants to calculate the F-statistic for each individual. Individuals whose F-statistic-based sex assignment was discordant with their reported sex at birth (F-statistic cutoff = 0.2) were removed.

For downstream XWAS, we retained biallelic SNPs with unambiguous alleles and MAF > 0.01, excluding the PARs as described above. Within each ancestry group, we assessed HWE among females and tested for differential MAF between sexes. We excluded variants that failed HWE in any ancestry group (p < 1e-15) or showed significant differential MAF between sexes in any ancestry group (p < 1e-7). The ancestry-specific QC failure lists were combined, and variants failing either criterion in any ancestry group were removed from the initial MAF-filtered variant set. After all QC, 365,774 chrX SNPs remained.

#### Sex-stratified XWAS

We performed pooled and sex-stratified XWAS with REGENIE [55] using age, age squared, and the first ten genetic PCs as covariates, as well as sex in the pooled analysis. After running the XWAS, we applied Bonferroni correction to adjust the p-values of SNPs. We used a chromosome X-wide significance threshold of p < 1.37e-7 (0.05/365,774), where 365,774 represents the largest number of variants tested across the three XWAS. We then obtained annotations and rsIDs in the same manner we obtained them for the autosomal SNPs.

### Sex differences in SNP effect sizes

To assess whether the effect sizes of genome-wide significant SNPs differed between women and men, we applied a z-score difference test to lead SNPs at distinct loci as determined by positional clumping of genome-wide significant variants from the combined GWAS. To identify lead variants, we selected all SNPs with a p-value below the significance threshold. We then performed distance-based clumping to retain only the most significant SNP within each 1,000 kb (1Mb) genomic window. Specifically, SNPs were sorted by ascending sex-combined p-value and the top SNP was designated as a lead variant. Any remaining SNP on the same chromosome whose physical position fell within 1,000 kb of that lead SNP was removed. We repeated this process iteratively until no candidate SNPs remained to be evaluated as potential lead SNPs.

Given the diversity of AoU participants with respect to genetic ancestry, we chose positional clumping as an ancestrally agnostic alternative to LD clumping, as a standard LD reference panel may not accurately reflect the LD patterns of this mixed ancestry group. This yielded 14 lead autosomal loci. For each lead SNP, we computed a z-score difference statistic:

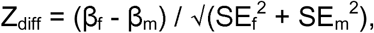

where β and SE denote the effect size and standard error from the sex-stratified GWAS. Two-sided p-values were calculated from the standard normal distribution and adjusted for multiple testing via Bonferroni correction across the 14 autosomal loci (0.05/14 = 3.57e-3). We applied the same correction to the SNP-by-sex interaction effects from the GxSex model at the same 14 lead loci. We assessed concordance between the sex-stratified and interaction analyses at the 14 lead loci by correlating the z-statistics.

### Heritability estimation

To estimate SNP-based heritability—the proportion of phenotypic variance in a population explained by additive genetic effects of common SNPs—we subset the analysis group to individuals identifying as “White” and “Not Hispanic or Latino” who were determined to be of European ancestry. We determined genetic ancestry using the AoU genetic similarity categories from the auxiliary file ancestry_preds.tsv, which groups individuals based on their genetic similarity to reference populations from the Genome Aggregation Database (gnomAD), Human Genome Diversity Project (HGDP), and 1000 Genomes. Specifically, we used the “1KGP-HGDP-EUR-like (EUR or European)” category to define this subset for heritability analyses. All heritability analyses are performed in the AoU Workbench using the genomic-relatedness-based restricted maximum-likelihood (GREML) approach from the Genome-wide Complex Trait Analysis (GCTA) toolkit [59]. We restricted our analysis to this group because GREML heritability estimation is biased in admixed populations [60] due to linkage disequilibrium differences across ancestral backgrounds. This subset represents the ancestry group in which GREML estimates are least susceptible to LD-induced bias, and is additionally the largest group in the AoU dataset.

We stratified this subset by sex and statin use status to produce four groups: untreated women, statin-treated women, untreated men, and statin-treated men. We estimate the SNP-based heritability of the unadjusted LDL-C phenotype in all groups as well as the statin-adjusted LDL-C phenotype in statin-treated individuals, using age, age squared, and the first ten genetic PCs in all models. We report heritability estimates for both the unadjusted and statin-adjusted LDL-C phenotypes in statin-treated individuals to provide a range that reflects uncertainty in the correction for statin effects, thereby providing a plausible interval within which the true additive genetic variance is likely bounded. The unadjusted phenotype is lowered by treatment, while the statin-adjusted phenotype assumes a uniform multiplicative effect that may not hold for all individuals.

To detect significant sex differences in SNP-based heritability estimates, we calculated a z-score difference statistic:

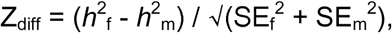

where *h*^2^ and SE denote the heritability estimate and standard error from the sex-stratified GREML results.

## Results

### Phenotypic analysis

#### Sex differences in median LDL-C levels

To evaluate potential sex-specific differences in LDL-C measurements in the combined analysis group (N = 86,700) comprised of women (n = 54,189) and men (n = 32,511) of diverse racial identities and predicted ancestries (Table S1), we calculated the Kullback-Leibler (KL) divergence, conducted a MWU test, and developed a HL estimator for both unadjusted and statin-adjusted LDL-C measurements. These analyses provide valuable insight into the distribution and central tendency of measurements of these two groups (Table S2 and Table S3, respectively).

In the unadjusted analysis (Figure 1a), the KL divergence between the LDL-C distributions of women and men was 0.05, indicating substantial overlap in distribution shape. The MWU test yielded a highly significant p-value (p < 2.2e-16), suggesting that the difference between groups lies in central tendency rather than distribution shape. The HL estimator, averaged over 1,000 bootstrap trials, quantified this difference at 11.42 mg/dL (95% CI: 11.00 to 12.00 mg/dL), indicating that the median LDL-C of women exceeded that of men by 11.42 mg/dL.

**Figure 1:**
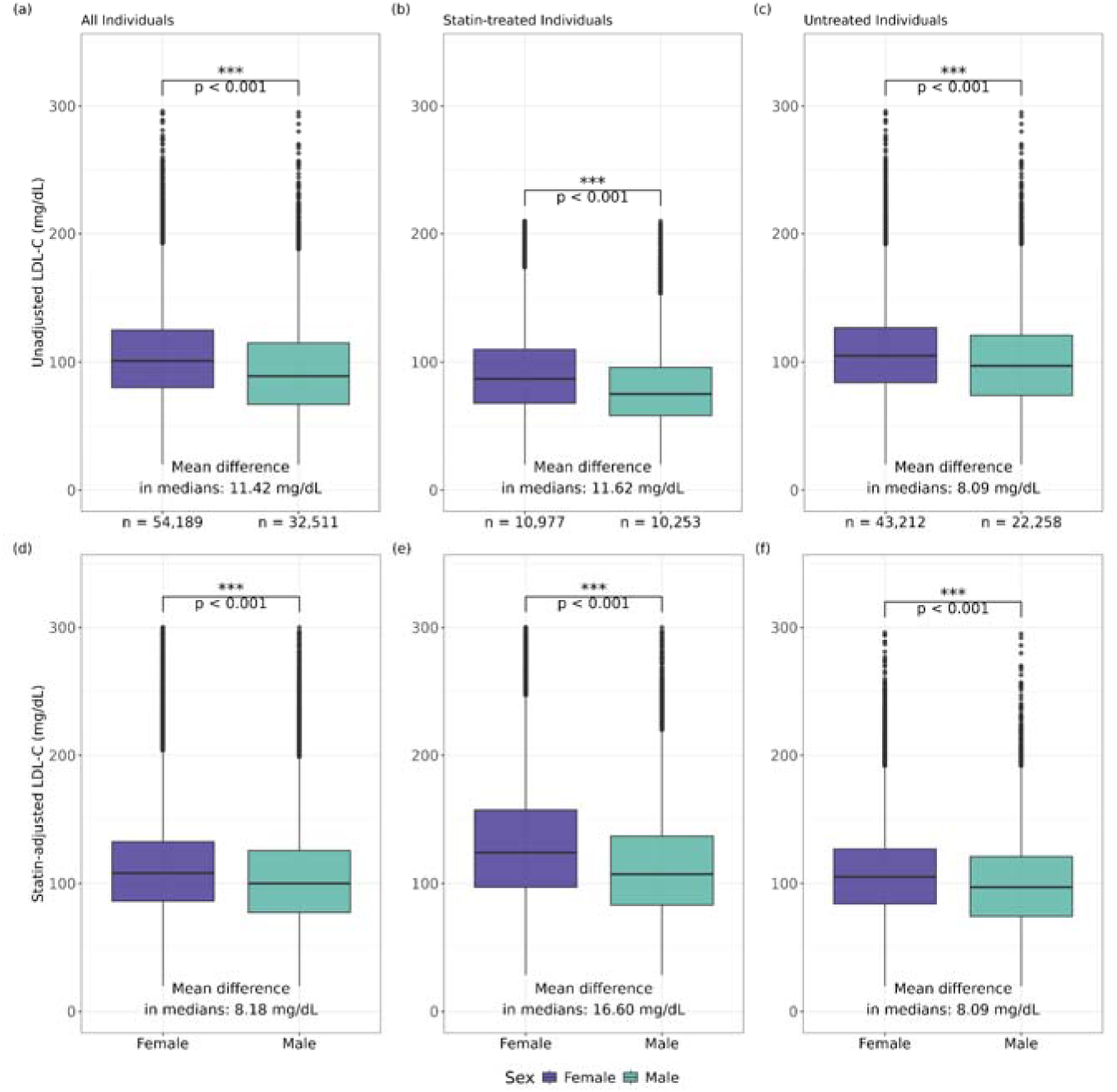
Statin-adjusted and unadjusted LDL-C levels by sex. Box plots of statin-adjusted (a–c) and unadjusted (d–f) LDL-C measurements (mg/dL) in women and men across three subgroups: (a, d) all individuals, women: n = 54,189; men: n = 32,511, (b, e) statin-treated individuals, women: n = 10,977; men: n = 10,253, and (c, f) untreated individuals, women: n = 43,212; men: n = 22,258. Boxes represent the interquartile range (IQR), with the horizontal line indicating the median. Whiskers extend to 1.5 x IQR; points beyond this range are plotted individually as outliers. The sex difference in medians as estimated by the Hodges-Lehmann estimator, averaged over 1,000 bootstrap trials, is annotated within each panel. Statistical significance was assessed using the Mann-Whitney U test; *** denotes p < 0.001. Across all three subgroups, LDL-C levels differed significantly between women and men.

In the statin-adjusted LDL-C analysis (Figure 1d), the KL divergence was 0.03 and the HL estimate was 8.18 mg/dL (95% CI: 8.00 to 9.00 mg/dL, p = 3.33e-227). The reduction in the HL estimate following adjustment suggests that a portion of the unadjusted sex difference in LDL-C in the combined group is attributable to the differential distribution of statin use between women and men. Nonetheless, a clinically meaningful difference persists after adjustment. An increase of 10 mg/dL in LDL-C is associated with a nearly 20% increase in heart attack risk [61]. Applying this estimate to our statin-adjusted analysis, the 8.18 mg/dL higher LDL-C observed in women corresponds to an approximately 16% higher relative risk of heart attack for women compared with men underscoring the significance of the residual difference observed here.

#### Sex-specific disparities in statin efficacy

To examine potential sex differences in statin efficacy, we restricted analyses to statin-treated individuals (women: n = 10,977; men: n = 10,253) using both unadjusted and statin-adjusted LDL-C measurements (Table S2, Table S3, respectively).

In the unadjusted statin analysis (Figure 1b), the KL divergence was 0.08, indicating somewhat greater distribution differences than in the combined group, though still reflecting substantial overlap. The MWU test again identified a significant difference in central tendency (p = 1.01e-167). The HL estimator averaged 11.62 mg/dL (95% CI: 11.00 to 12.00 mg/dL), indicating that statin-treated women had a higher median LDL-C than statin-treated men by 11.62 mg/dL. In the statin-adjusted analysis (Figure 1e), this difference increased to 16.60 mg/dL (95% CI: 15.71 to 17.14 mg/dL, p = 1.01e-167), while the KL divergence remained 0.08.

The stability of the distribution shape alongside the increase in the HL estimate after adjustment indicates that adjustment reveals a larger underlying difference in central tendency than the raw measurement suggests. This pattern is consistent with statins more effectively lowering LDL-C in men than in women within this cohort, such that unadjusted measurements may underestimate any true sex difference in statin response.

These findings are consistent with lower efficacy of statins in women as compared to men, underdosing of statins in women, or both. Given that women in this study generally have higher LDL-C measurements than men, it is possible that current dosing schemes are not adequate for women [62]. This association may reflect gender differences at the clinical level that could lead providers to underdose women with statins, as well as possible sexually dimorphic effects that may reduce the efficacy of statin medications even were they appropriately dosed. It is also worth noting that most major statin trials and clinical dosing guidelines have been developed from cohorts composed primarily of men [62], which may contribute to differential treatment outcomes observed between sexes.

#### Sex differences in LDL-C among untreated individuals

We repeated these analyses in individuals not receiving statin therapy (women: n = 43,212; men: n = 22,258; Figure 1c, Figure 1f). The KL divergence was 0.03 in both the unadjusted and statin-adjusted analyses, as expected given that adjustment does not affect untreated individuals. The HL estimate was 8.09 mg/dL (95% CI: 8.00 to 9.00 mg/dL, p = 2.82e-182) in both analyses, indicating that the median LDL-C of women exceeded that of men by approximately 8.09 mg/dL independent of statin use. This residual difference in the untreated group suggests an underlying sex difference in LDL-C that is not attributable to differential statin exposure.

#### Sex differences, and similarities, in maximum LDL-C

Maximum LDL-C presents a more complex picture than above (Figure S1, Figure S2, Table S4). In statin-treated individuals, the statin-adjusted HL estimate for maximum LDL-C in the 20–300 mg/dL range was 20.28 mg/dL (p = 1.7e-97)—larger than the primary analysis, consistent with maximum LDL-C better capturing pre-treatment values. However, in untreated individuals the statin-adjusted HL estimate was 1.02 mg/dL (p = 1.1e-3), and the statin-adjusted all individuals result showed a small difference in the opposite direction (HL = -0.83 mg/dL, p = 0.013), both driven by a near-null sex difference in the untreated group. The sex difference in maximum LDL-C appears to be primarily driven by the statin-treated group, underscoring the importance of accounting for statin use when estimating sex-specific LDL-C differences.

#### Most recent and maximum LDL-C sensitivity analyses

We conducted sensitivity analysis comparing the primary phenotype we analyzed (most recent LDL-C, filtered to 20-300 mg/dL) to maximum LDL-C across three filtration ranges: <1,000 mg/dL, 10--500 mg/dL, and the primary 20-300 mg/dL range. We examined sex differences in unadjusted and statin-adjusted LDL-C across all individuals, statin-treated individuals, and untreated individuals. To enable direct comparison, we also examined most recent LDL-C under the same filtration ranges for both statin-adjusted and unadjusted LDL-C (Tables S5 and S6, respectively).

Most recent LDL-C was largely stable across filtration choices. In untreated individuals, the statin-adjusted HL estimate was 8.16, 8.16, and 8.09 mg/dL across the filtration ranges, respectively. The statin-treated HL estimates were similarly consistent at 17.38, 17.35, and 16.60 mg/dL.

Maximum LDL-C presented a more complex pattern. The statin-treated results were consistent with the primary analysis and robust across ranges, with statin-adjusted HL estimates of 23.99, 23.86, and 20.28 mg/dL—larger than the primary analysis, consistent with maximum LDL-C in statin-treated individuals better capturing pre-treatment values. However, untreated individuals showed little sex difference regardless of filtration choice (HL ≈ 1.02mg/dL across all three ranges), compared to approximately 8 mg/dL in the analyses using most recent LDL-C. The statin-adjusted all-individuals result is also not significant in the <1,000 mg/dL and 10–500 mg/dL ranges (p = 0.088 and p = 0.076, respectively), driven by the untreated group.

The distributional shape of the untreated maximum LDL-C was more divergent between sexes in the <1,000 mg/dL range (KL divergence ≈ 1.9 vs. 0.003–0.004 in capped versions), suggesting that the choice of phenotypic range substantially changes the composition of the sample. In contrast, the most recent LDL-C phenotype produced consistent, stable results across all filtration ranges, while maximum LDL-C results varied across treatment strata and were more sensitive to filtration decisions—particularly in the untreated group, where the sex differences essentially disappeared.

#### Sex differences in statin treatment rates

To assess how treatment rates may differ between sexes, we stratified women and men into the following age groups by decades: 21–30, 31–40, 41–50, 51–60, 61–70, and 71–80. We then calculated the median LDL-C value for each sex and age group combination (Tables S7 and S8). For statin-treated individuals, we also calculated a statin treatment rate, calculated as the number of statin-treated individuals within each sex and age group divided by the total number of individuals in that sex and age group. With the exception of the 21–30 age range, women consistently have the highest median LDL-C measurements. Despite this, men are treated with statins at a higher rate than women across all age strata (Figure 2a). This result is consistent with men receiving treatment earlier and more aggressively than women for LDL-C level management. Furthermore, our results are consistent with statins being less effective at lowering LDL-C in women than in men, as indicated by women retaining the highest median levels even among the statin-treated group.

**Figure 2:**
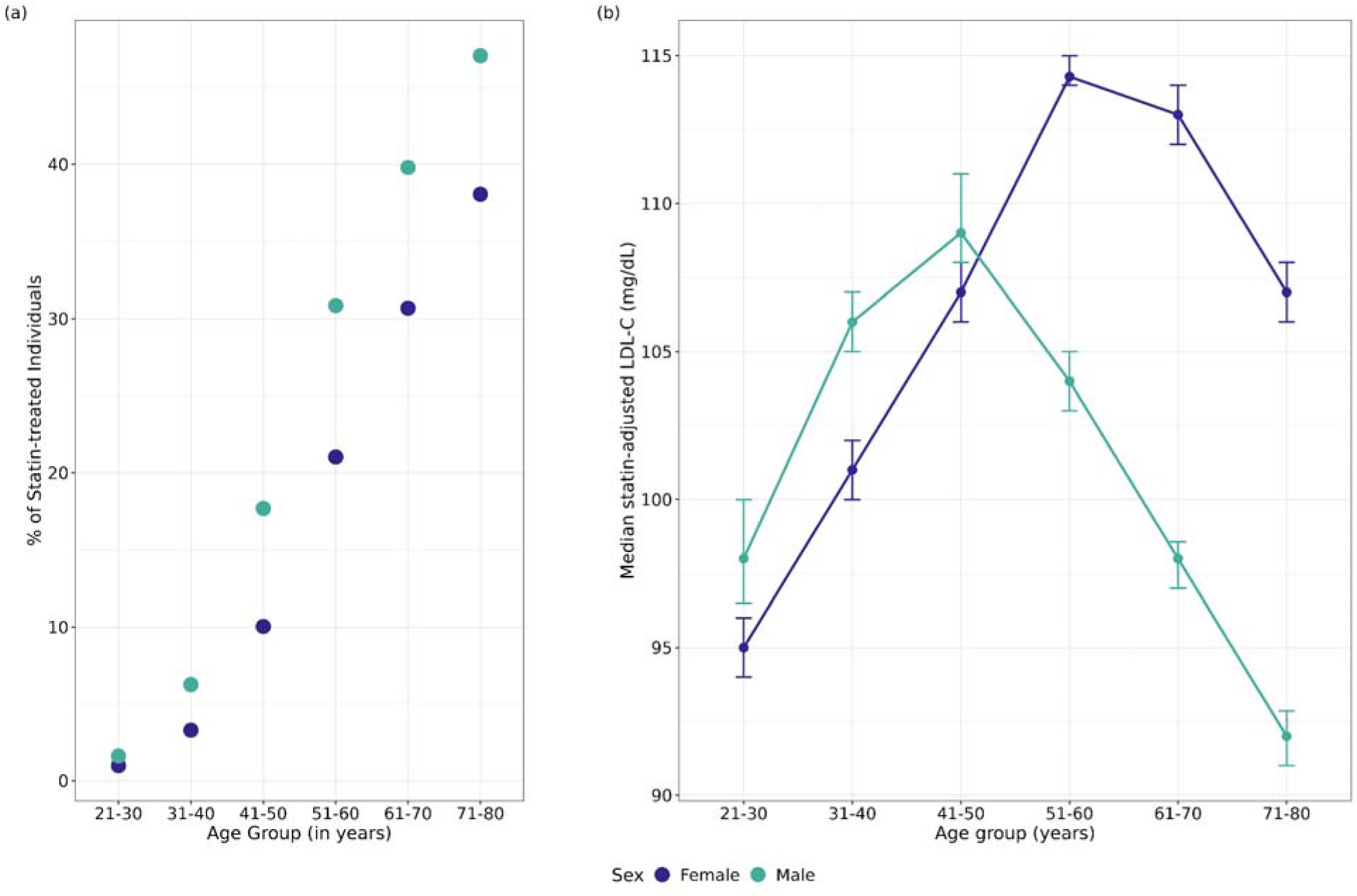
Sex differences in statin treatment rates and statin-adjusted LDL-C distributions across age groups. (a) Scatterplot of statin treatment rates in women (purple) and men (green) by age group, calculated as the number of statin-treated individuals divided by the total number of individuals of each sex within each age group. Statin treatment rates increase with age in both sexes, with higher rates among men across all age groups except ages 21–30. (b) Median statin-adjusted LDL-C levels by sex and age group, with error bars indicating the 95% confidence interval around median estimates. Median LDL-C levels increase through midlife in both sexes but diverge after ages 41–50. In women, median LDL-C peaks at ages 51–60, whereas in men it declines after ages 41–50.

#### Sex-specific age-related changes in LDL-C

To assess how LDL-C measurements vary across ages, we plotted the median statin-adjusted LDL-C measurements for each age stratum (Figure 2b). In both sexes, there is an increase of the median of statin-adjusted LDL-C that corresponds with age. The shift is more apparent in the age groups that correlate with peri-menopause and menopause, 41–50 and 51–60, followed by a decline in the 61–70 and 71–80 strata. Research has established that LDL-C increases with the loss of estrogen [63] [64]. In contrast, the median LDL-C measurements for men remain relatively stable and shift lower with age.

### Phenotypic analysis, racially stratified

#### Racial differences in LDL-C levels and statin treatment efficacy

To examine whether sex-specific differences varied by race, we stratified the combined analysis group both by sex and the two self-reported racial identities with the largest sample size—“White” and “Black or African American”. We calculated the median LDL-C values for individuals on and off statin therapy within each subgroup and conducted MWU-tests to assess statistical significance. We note that sample sizes varied considerably across subgroups, with White individuals outnumbering Black individuals approximately 3–4 fold among untreated individuals and among statin-treated individuals (Table S9). Given the 3- to 4- fold smaller sample sizes of Black individuals, these stratified estimates have reduced precision; accordingly, the observed racial patterns should be considered exploratory and hypothesis-generating as they are insufficient to support robust conclusions about racial differences in LDL-C levels or statin-efficacy.

Among untreated individuals, median LDL-C levels were lower in Black women (101 mg/dL) than White women (106 mg/dL), and in Black men (94 mg/dL) than in White men (97 mg/dL), though differences were small (Figure S3, Table S9). However, statin-treated Black women had the highest median LDL-C levels compared to any other subgroup, and higher median LDL-C levels (127 mg/dL) compared to statin-treated White women (123 mg/dL). This pattern was even more pronounced in men, with statin-treated Black men showing higher median LDL-C levels (116 mg/dL) than statin-treated White men (106 mg/dL). These findings suggest that statin efficacy may differ not only by sex but also by racial identity, potentially due to prescribing patterns and access.

### GWAS

#### Validation of combined analysis group GWAS

We performed GWAS on the combined analysis group and sex-stratified subgroups including age, age squared, and the first ten genetic PCs as covariates. In addition, the combined analysis group GWAS (Figure S4) included sex as a covariate. We compared linear models with and without LDL-C measurement method using the same GWAS phenotype and covariates. Measurement method explained only 0.35% of the residual variance in statin-adjusted LDL-C after accounting for all other covariates (partial R^2^ = 0.0035), and the total model R^2^ increased from 2.93% to 3.26% when method was added—an incremental gain of 0.33% (Table S10). No major sex differences were observed in the distribution of measurement methods (Table S11). Given the minimal contribution of measurement method to residual variance, we did not include it as a covariate in the GWAS models.

The combined analysis group reproduced known results and had an acceptable genomic inflation factor (λ = 1.06) (Figure S5). The combined analysis group GWAS effect sizes were in strong agreement (R^2^ = 0.927, p = 2.8e-12) with the TOPMed LDL-C effect sizes of significant hits GWAS (Figure S6), which was performed in a dataset of participants of diverse ancestries [23]. Importantly, we note that the TOPMed GWAS uses a rank-inverse normal transformed phenotype while our study uses an untransformed LDL-C in mg/dL; the strong agreement in effect sizes at shared genome-wide significant loci therefore holds across this methodological difference providing additional confidence in the robustness of our results.

#### Sex-stratified genetic associations for LDL-C

Using REGENIE [55], we ran GWAS on the two independent groups of women and men. We ran these analyses with age, age squared, and the first ten genetic PCs as covariates (Figure 3a). The sex-stratified analysis reproduced known results and had acceptable genomic inflation factors (λ ≤ 1.05) (Figure S7). After Bonferroni test correction, the GWAS identified 169 genome-wide significant SNPs in women (Supplementary Data 1a), 115 of which were significant in women only, and 55 genome-wide SNPs in men (Supplementary Data 1b), one of which was significant in men only. Genome-wide significant SNPs from the combined analysis are reported in Supplementary Data 1c. Of these sex-stratified results, two SNPs identified in women were not significant in the GWAS of the entire combined analysis group; however, all SNPs identified in men were also significant in the combined analysis group GWAS. Given the larger sample size of women (n = 54,189) than of men (n = 32,511), the greater number of SNPs reaching significance in women only may partly reflect increased statistical power to detect smaller effects.

**Figure 3:**
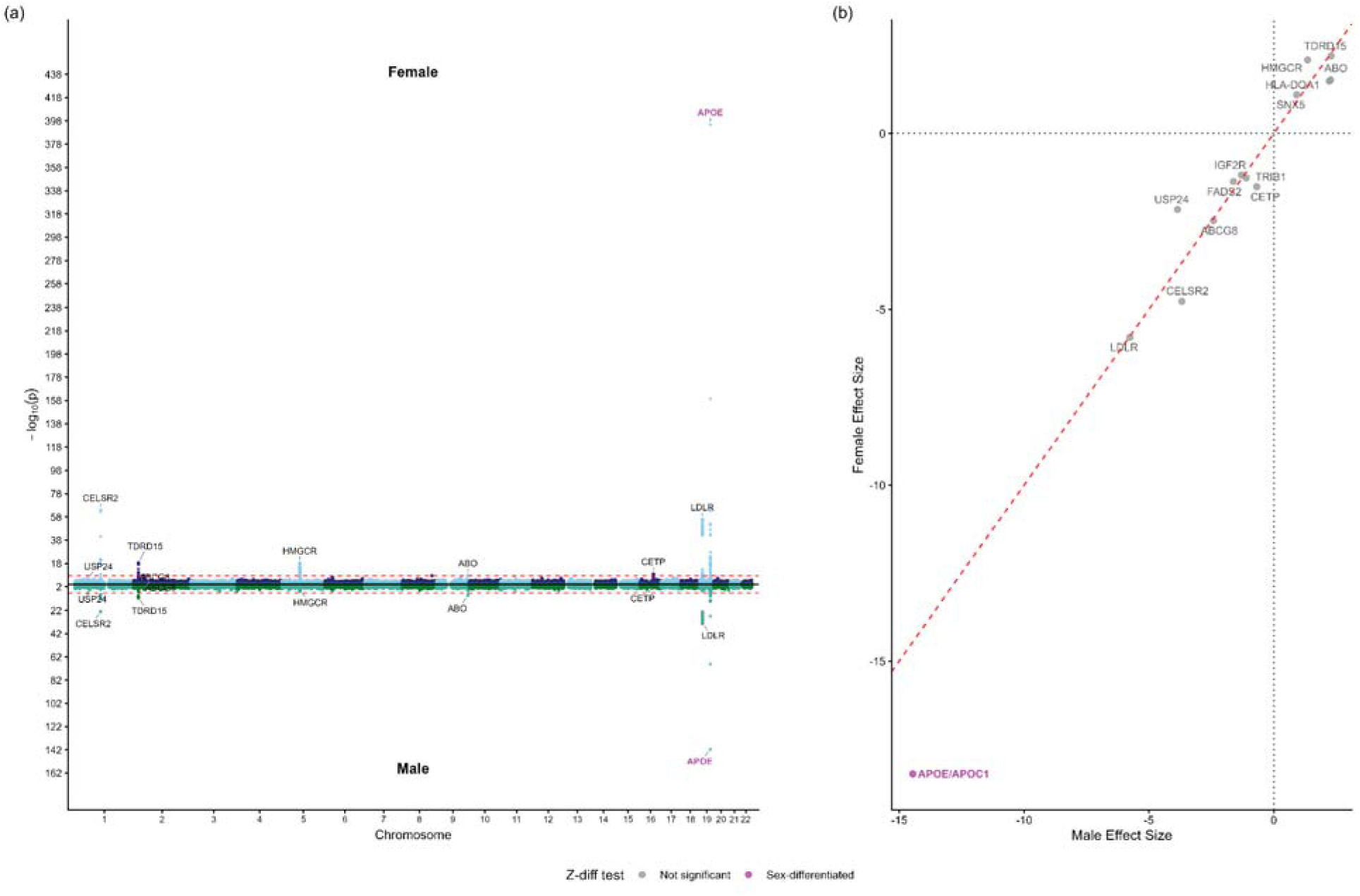
Sex-stratified GWAS of statin-adjusted LDL-C and sex differences in lead SNP effect sizes. (a) Miami plot of GWAS results for 54,19 females (top, points above zero) and 32,511 males (bottom, points below zero) for statin-adjusted LDL-C. Red dashed lines indicate the Bonferroni-corrected genome-wide significance threshold. Gene names bolded in pink denote loci with statistically significant sex-differentiated effects based on z-score difference test. (b) Scatterplot comparing effect size estimates from the female GWAS (y-axis) and male GWAS (x-axis) for the 14 lead loci. The red dashed diagonal indicates equal effect sizes between sexes; points are colors by z-score difference test significance (gray, not significant; pink, sex-differentiated). The *APOE/APOC1* locus showed a statistically significant sex-differentiated effect (Z_diff_ = -5.28, p = 1.78e-6), with a larger LDL-C lowering effect in women (β = -18.22 mg/dL) than in men (β = -14.45 mg/dL). The remaining 13 loci showed no significant sex-differentiated effects.

#### Sex differences in genetic effect sizes

We assessed whether effect sizes at genome-wide significant loci differed between women and men using a z-score difference test applied to 14 lead SNPs identified by positional clumping of combined GWAS results (Figure 3b, Table S12). Of the 14 loci tested, one—the *APOE/APOC1* locus on chromosome 19 (rs3200542)—showed a statistically significant sex-differentiated effect after Bonferroni correction (Z_diff_ = -5.28, adjusted p-value = 1.78e-6). The effect on LDL-C was larger in women (β = -18.22 mg/dL, SE = 0.428) than in men (β = -14.46 mg/dL, SE = 0.569). Both *APOC1* and *APOE* have established roles in lipid metabolism [65], [66] and *APOC1* has been implicated in immune and inflammatory responses and diabetes [66], [67], [68]. This region is characterized by strong LD between *APOE* and *APOC1*; therefore, without fine-mapping or conditional analyses, we cannot determine whether this signal represents an independent *APOC1* effect or tagging *APOE* haplotypes. Nonetheless, our finding of a sex-differentiated effect at this locus is consistent with prior studies reporting sex-based variation in the *APOE/APOC1* region [20], [69]. The remaining 13 loci showed no significant sex-differentiated effects. Effect-size differences at *CETP* and *HMGCR* were in the same direction of previous analyses, larger in women [20], while the nominal direction of the sex difference at *CELSR2* was opposite to previous analyses [20]. However, these loci did not demonstrate significant sex-differentiated effects in our analyses, potentially due to statistical power.

#### GxSex interaction

In the genome-wide GxSex scan of the combined analysis group, no variant reached genome-wide significance (p < 4.94e-8, Figures S8 and S9). The strongest signal was at the *APOE/APOC1* locus where the SNP-by-sex term was β = 3.54 (SE = 0.657, p = 7.19e-8), just under the genome-wide significance threshold. With females as the reference, the positive interaction coefficient indicates that the LDL-C lowering effect of this locus is stronger in women than in men, consistent with the sex-stratified estimates (women β = -18.22; men β = -14.46).

At the 14 lead loci identified in the combined sex GWAS (which did not include an interaction term) the SNP-by-sex interaction results agreed with the sex-stratified z-score difference test described above (Table S13): *APOE/APOC1* was the only locus significant after correction and no other locus was significant after correction. The z-statistics from the two approaches were highly correlated across the 14 lead loci (r = 0.970, including *APOE/APOC1*). Though *APOE/APOC1* showed a significant sex difference based on the locus-level analysis under both approaches (Figure S10), it did not reach significance in the genome-wide GxSex scan.

### XWAS

#### Sex-stratified chrX associations for LDL-C

We performed XWAS on the combined analysis group as well as the two independent groups of women and men using the same covariates as the autosomal GWAS and statin-adjusted LDL-C as the phenotype (Figure 4). All three analyses had acceptable genomic inflation factors (λ ≤ 1.09) (Figure S11). After Bonferroni test correction, no significant SNPs were detected in the combined analysis group or female restricted analyses. One SNP, rs73461969, reached X-wide significance only in men, but not in women or the combined analysis group (p = 3.32e-8, 0.96, and 1.47e-4, respectively). This SNP is within the *PTCHD1-AS* region of the X chromosome, a gene that has previously been associated with autism spectrum disorders in men [70] and with LDL-C [71]. Because rs73461969 was significant in men only, we did not test for a sex difference in effect size.

**Figure 4:**
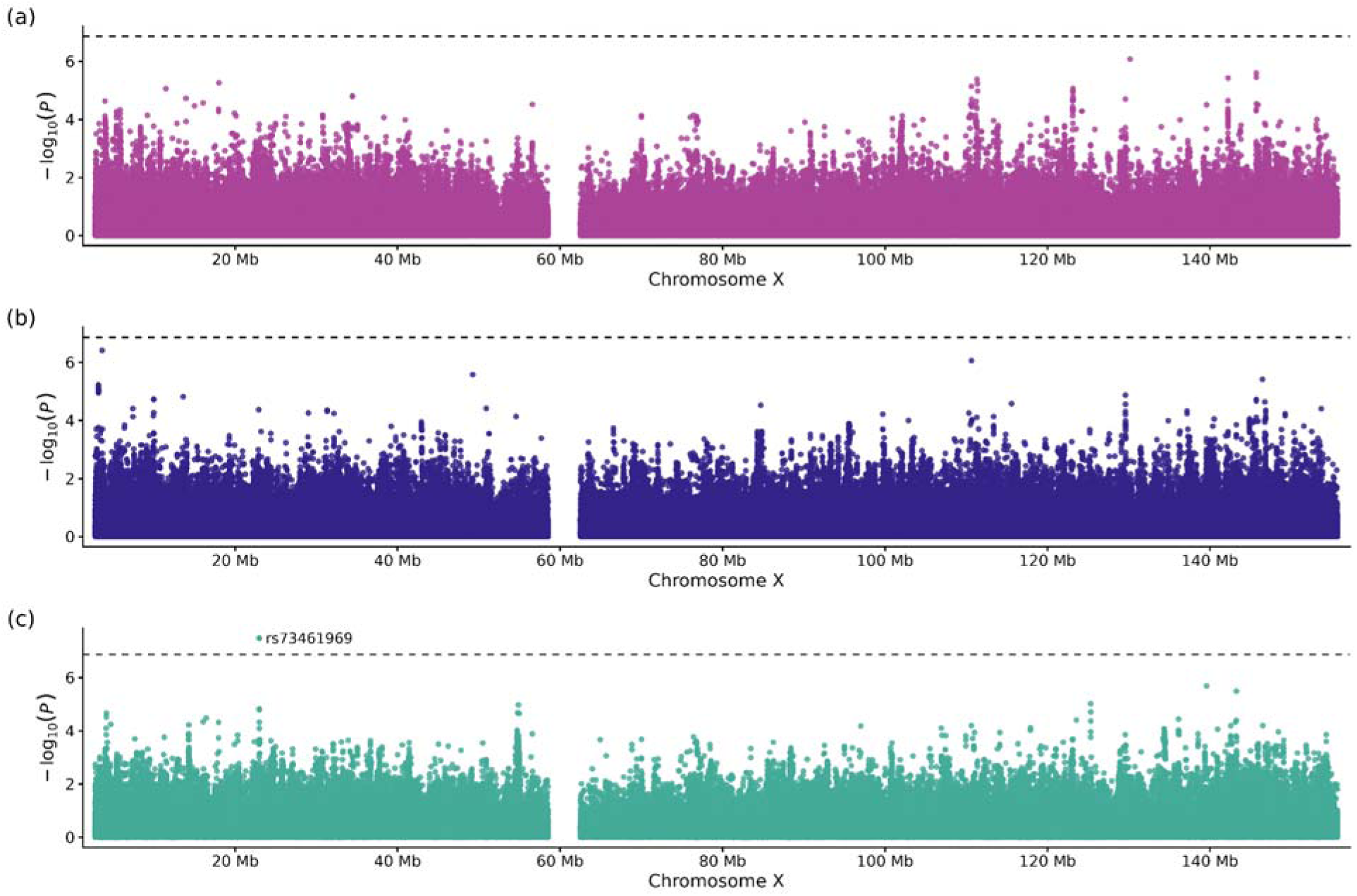
Manhattan plots of XWAS of LDL-C. Plots display SNP position on the x-axis and -log_10_(p) values on the y-axis for (a) the combined analysis group (pink), (b) women only analysis (purple), and (c) men only analysis (green). No significant associated loci are identified in the pooled or women only analyses. The SNP, rs73461969 within the *PTCHD1-AS* region, was statistically significant in men only.

Three variants previously reported in sex-stratified XWAS of LDL-C reached nominal significance (p < 0.05) in our combined analysis group including rs1802288 in the *TSPAN6* region, rs1050828 in the *G6PD* region, and rs2519273586 in the *ASB9/ASB11* region; rs2519273586 was also nominally significant in men; an additional previously reported variant, rs7886627 in the *FAM50A/PLXNA3* region, reached nominal significance in men [20]. None survived test correction.

### Heritability

#### Most recent LDL-C measurement

We assessed whether SNP-based heritability estimates (*h*^2^, which is calculated as V(G)/Vp) differed between women and men. To do so, we applied a z-score difference test to each combination of statin treatment status and LDL-C phenotype (unadjusted for statin use and adjusted for statin use) (Figure 5, Figure S12, Table S14). This analysis included untreated women (n = 22,829), statin-treated women (n = 5,796), untreated men (n = 12,830), and statin-treated men (n = 6,289). Of the three combinations tested, none showed significant differences between sexes. Across the LDL-C-by-statin strata, there is no statistical evidence that *h*^2^ differs by sex. Furthermore, no significant sex differences were found in genetic variance (V(G)), suggesting that any sex differences in additive genetic contribution to LDL-C variation, if present, are not large enough to detect in these strata. However, power to detect sex differences in SNP-based heritability and genetic variance was limited in the statin-users due to smaller sample sizes and large standard errors.

**Figure 5:**
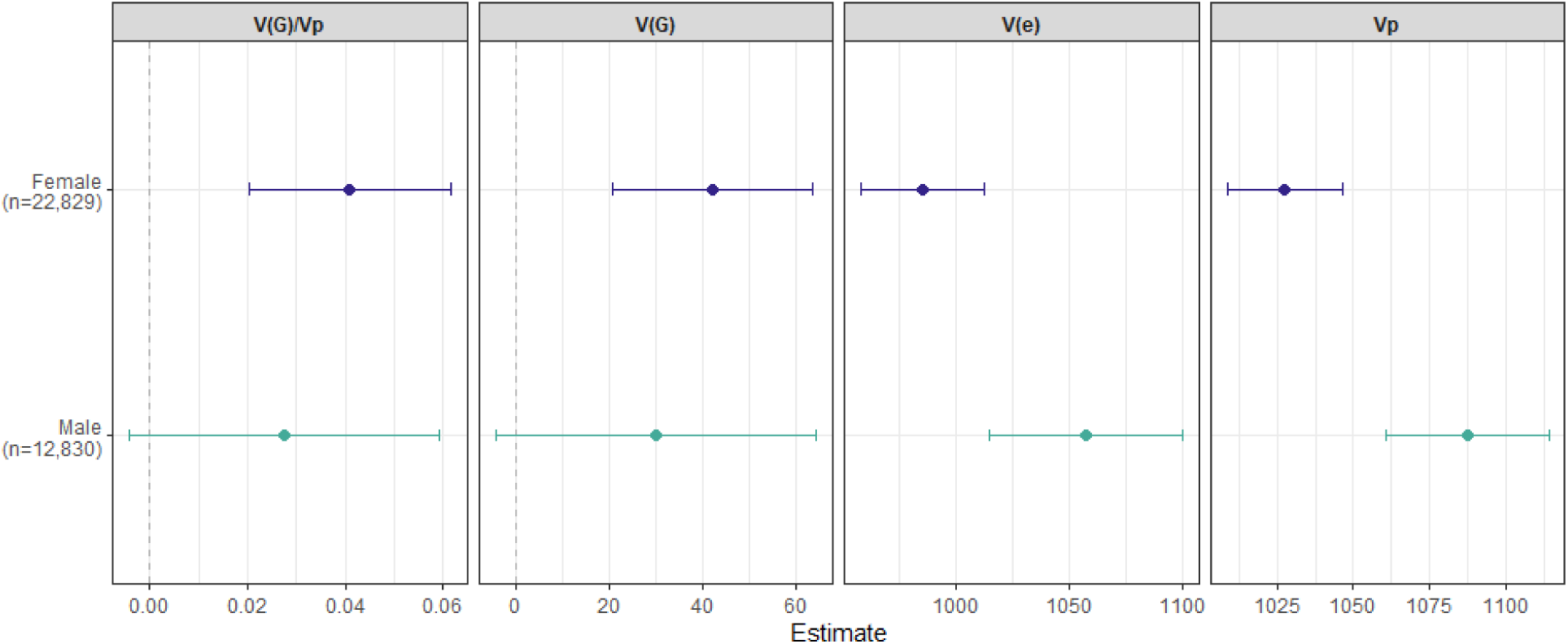
SNP-based heritability and variance components by sex (untreated individuals). Forest plot showing the SNP-based heritability (leftmost panel V(G)/Vp), genetic variance (second panel, V(G)), residual variance (third panel, V(e)), and phenotypic variance (rightmost panel, Vp) estimates for untreated women (purple) and men (green). Heritability and additive genetic variance do not differ significantly between women and men as confirmed by a z-score difference test. However, V(e) is significantly (Z_diff_ = -2.79, p-value = 0.005) lower in women (V(e) = 985) vs men (V(e) = 1057). Additionally, Vp differs between sexes, being lower in women (Vp = 1028) than in men (Vp = 1087).

In contrast, residual variance (V(e)) showed significant sex differences in all strata. In the untreated individuals (Figure 5), V(e) was significantly (Z_diff_ = -2.79, p-value = 0.005) lower in women (V(e) = 985) vs men (V(e) = 1057). In the statin-treated individuals, for both the statin-adjusted (Figure S12a) and non-adjusted (Figure S12b) phenotypes, V(e) was significantly (Z_diff_ = 3.07, p-value = 0.002) higher in women (statin-adjusted V(e) =1884, unadjusted V(e) = 923) than in men (statin-adjusted V(e) = 1580, unadjusted V(e) = 774).

Phenotypic variance (Vp) followed a similar pattern with significant sex differences across all groups (all p < 0.001). The direction mirrored V(e), lower in women not using statins (Vp = 1028) than in men not using statins (Vp = 1087). In statin-treated individuals, for both phenotypes, Vp was higher in women (statin-adjusted Vp = 1957, unadjusted Vp = 959) than in men (statin-adjusted Vp = 1649, unadjusted Vp = 808). While the sex differences in residual variance as well as phenotypic variance are statistically robust, the magnitude of V(e) and Vp in statin-treated individuals should be interpreted with caution because the adjustment for statin use assumes a uniform multiplicative effect that may not fully account for sex-specific statin response or adherence.

The magnitude of the difference in V(e) and Vp between the statin-adjusted and unadjusted phenotypes in statin-treated individuals is an expected consequence of linear rescaling. The ∼ 2.04 ratios for the variance components between the statin-adjusted and unadjusted phenotypes in statin-treated individuals are the direct mathematical consequence of using the standard adjustment of 1/0.7 (since (Var(*c*X) = *c^2^* Var(X)), which fully accounts for the observed magnitude differences in the variance components while leaving heritability invariant.

These results indicate that while genetic architecture of LDL-C, as measured by SNP-based heritability, does not show detectable differences between sexes in these strata, environmental or residual influences contribute differentially to trait variance in females versus in males.

#### Maximum LDL-C measurement

In the supplementary analyses using maximum LDL-C in the primary range of 20–300 mg/dL as the alternative phenotype (Figure S13, Table S15; untreated women n = 22,541, statin-treated women n = 3,996, untreated men n = 12,840, statin-treated men n = 4,620), SNP-based heritability (*h*^2^, as V(G)/Vp)) and additive genetic variance (V(G)) largely paralleled the primary findings in most recent LDL-C and showed no significant sex differences across any stratum (all p ≥ 0.467), reinforcing the null results for additive genetic contributions. For residual variance (V(e)) and phenotypic variance (Vp), the non-statin stratum (Figure S13a) again demonstrated significantly lower values in women than in men (V(e): Z_diff_ = -4.45, p-value < 0.001; Vp: Z_diff_ = -7.21, p-value < 0.001), consistent with the pattern observed for most recent LDL-C.

In contrast to the primary analysis, neither V(e) nor Vp showed significant sex differences within statin-treated individuals, regardless of phenotype adjustment (all p ≥ 0.170) (Figure S13b and Figure S13c). Notably, variance estimates in the statin-treated groups carried large standard errors, indicating statistical uncertainty in these estimates.

These results support the conclusion that sex differences in additive genetic effects are undetectable for maximum LDL-C, while underscoring that sex-specific residual contributions to trait variance are less consistently observed in statin-treated individuals.

## Discussion

In this study, we report sex-specific differences in LDL-C phenotype, management, treatment, and largely concordant genetic architecture. We found that women in this data set have higher median LDL-C levels than men, receive statins less often, and seem to respond to them less effectively. Women consistently demonstrate higher LDL-C measurements as compared to men across most age strata. This difference widens with age, with women experiencing an LDL-C level increase in age ranges that correlate with the onset of peri-menopause and menopause (40 to 60) and the current recommended age range to begin routine lipid screening (beginning at 45).

For the maximum LDL-C measurements, the pattern we observed differed from the primary analysis. In sensitivity analyses comparing most recent and maximum LDL-C across three filtration ranges, most recent LDL-C remained stable across filtration choices. In contrast, maximum LDL-C results were treatment strata dependent and sensitive to filtration ranges. One possible explanation—though not directly tested here—is that women and men share a similar biological upper limit, or ceiling, for LDL-C even if their baseline levels differ, in which case max LDL-C would capture a sex-invariant ceiling effect rather than the baseline our study aims to characterize. This interpretation is consistent with the most recent LDL-C showing higher medians in women across all subgroups, and with the statin-treated individuals max LDL-C results, which reinforce our primary findings regarding sex differences in statin-treated LDL-C levels and sex-differentiated statin effects.

An alternative, non-competing explanation is that maximum LDL-C, by capturing each individual’s recorded highest value rather than a temporally matched measurement, may combine peak values from different stages of life. We’ve shown that median LDL-C in women rises through the age strata corresponding to peri-menopause and menopause, peaking in the 51–60 stratum, and subsequently declining in the 61–70 and 71–80 groups; in men, median LDL-C peaks earlier, in the 41–50 stratum, and declines continuously thereafter. Because women’s LDL-C peak occurs roughly a decade later than men’s, if the untreated subgroup spans a range of ages relative to menopause, the max-based phenotype may hide a sex difference whereas the most recent phenotype may not. This candidate explanation is consistent with the age-related widening of the sex difference we describe above, though we did not directly test it here.

Whether the null median difference in untreated individuals reflects an artifact of combining peak values from individuals at different ages or menopause stages, true biological similarity, measurement error, or some combination of these factors remains unclear from these data alone.

In the absence of statin therapy, racial differences in median LDL-C were minimal. Among statin-treated individuals, however, Black participants exhibited higher on-treatment LDL-C levels than their White counterparts, with Black women showing the highest absolute LDL-C levels of any subgroup and the racial gap itself being somewhat larger among men. This divergence suggests these observed disparities are not attributable to baseline biological differences, but rather emerge in the context of statin treatment. This may reflect systemic inequities in access, as prior studies have shown Black women have lower odds of receiving statin therapy compared to White women even when clinically indicated [72]. The pattern of Black individuals having higher median LDL-C levels despite statin treatment compared to their White counterparts is consistent with previously reported differences in statin response across racial and sex groups [73], [74], [75], [76], [77].

Our sex-stratified GWAS also identified sex-differentiated effects of *APOE/APOC1*, with a larger LDL-C lowering effect in women than in men. This finding is consistent with prior reports of sex-based variation at the *APOE/APOC1* locus, and given the well-established role of *APOE* in lipid metabolism, it raises the possibility that at least part of the sex difference in LDL-C has a genetic basis at this locus. The sex difference at this locus was confirmed in a locus-specific GxSex interaction model, but no variant, including *APOE/APOC1*, reached genome-wide significance in the interaction scan. This could be because interaction tests have lower power than corresponding main-effect tests: the standard error of a sex difference is larger than that of either stratified estimate, so this result is expected. We also observed an asymmetry in the number of genome-wide significant SNPs identified in each sex-stratified GWAS: 169 in women (115 of which were significant in women only) compared with 55 in men (only one of which was significant in men only). Given the smaller sample size in men (n = 32,511) compared to women (n = 54,189), the power to detect small but true sex-differentiated effects in our study is limited. The observed difference at the *APOE/APOC1* locus, while statistically robust, may still be influenced by differences in sample size and precision between the two groups and larger, well-powered studies are needed to confirm whether additional loci exhibit true sex-specific effects on LDL-C.

Our pooled and sex-stratified XWAS were unable to recapitulate many known LDL-C associations on chrX [20], potentially due to statistical power. The X-wide significant SNP in men lies in the *PTCHD1-AS* region, a gene that has previously been associated with autism spectrum disorders in men [70] as well as with LDL-C [71]. This is consistent with evidence of overlap between autism spectrum disorders and cardiometabolic traits [78], including changes in lipid metabolism and LDL-C specifically [79]. Given the sex-specific biology, this region could inform future work on the shared genetic architecture of autism spectrum disorders and lipid traits.

Heritability and additive genetic variance for LDL-C did not differ significantly between sexes in any stratum (though power was limited in statin-treated individuals) suggesting that the additive genetic contribution of common SNPs is similar in women and men. These findings were consistent when using maximum LDL-C as the alternative phenotype, restricted to the primary 20–300 mg/dL filtration range. However, residual variance showed robust sex differences, with lower variance in non-statin using women and higher variance in statin using women, pointing to sex-specific non-genetic influences on LDL-C variability. Consistent with this, a recent study decomposing sex differences in trait correlations in the UK Biobank found that environmental—rather than genetic—factors were the primary contributors to sexual dimorphism [80]. For maximum LDL-C, this pattern was observed in untreated individuals, but not in statin-treated individuals, where variance estimates had large standard errors, indicating statistical uncertainty. Despite this, the ratio of statin-adjusted to unadjusted variance estimates in statin-treated individuals closely matched the theoretical expectation of 1/0.7^2^ ≈ 2.04 under linear rescaling for both most recent and maximum LDL-C, confirming that this scaling relationship holds even where the absolute magnitude of the underlying variance estimates is poorly determined.

A more relevant concern in our study is the comparison between maximum and most recent LDL-C. Unlike the statin adjustment, which is a known linear transformation of a single underlying phenotype, maximum and most recent LDL-C are not scale transformations of one another, but rather distinct derived phenotypes, representing different ways of capturing an individual’s LDL-C measurements from EHR data, and their differing sensitivity to filtration choices reflects that variance-based estimates in genetic analyses can be sensitive to how a phenotype is defined, independent of any true underlying differences in variability. This distinction is relevant to genetic studies that use EHR measurements as their primary phenotype, where the choice of which phenotype to use—rather than any subsequent linear adjustment—may be the more consequential source of imprecision.

This recapitulates our phenotypic findings above, where most recent LDL-C produced stable sex differences across all filtration choices while maximum LDL-C results varied by treatment strata and filtration range. Taken together, these observations suggest that the choice of phenotype may be a contributing factor to the statistical uncertainty we observed in the variance estimates derived from these two phenotype definitions. However, this is likely compounded by the limited sample sizes in the statin-user strata making it difficult to disentangle the effect of phenotype definition from the effect of statistical power. Therefore, while phenotype definition is an important consideration, we cannot attribute the imprecision solely to that choice. Overall, these heritability analyses indicate that any sex differences in LDL-C levels are not explained by differential additive genetic architecture in this group. Instead, environmental or residual factors—including potentially sex-specific measurement error, lifestyle, or unmodeled biological responses (such as sex-hormone signaling, gene-by-environment interactions, or sex-specific statin responses)—may contribute to the greater total variance observed in untreated men and statin-treated women. This reinforces the importance of considering sex in clinical management where environmental or residual exposures may differ systematically.

These findings underscore the importance of considering sex-specific approaches when examining lipid profiles and treatment options. Addressing these differences may include the establishment of different clinically relevant LDL-C thresholds for women and men and the development of alternative pharmaceutical solutions that have improved efficacy in women. Our findings suggest that current approaches may not be equitable across the sexes, and clinical care guidelines may need to consider evolving evidence to ensure that CVD risk is effectively managed in each group.

Our study has several limitations related to data sources, cohort composition, and model scope. First, sex, gender, race, and ethnicity were all obtained through self-report survey data. While these social identifiers are important, electing to use self-report data, a single biobank, and narrow definitions of sex could potentially limit the interpretation and transferability of our results. Second, we note that the statin adjustment factor of 0.7 may not be equally accurate for women and men as a biological correction for statin efficacy; this represents a limitation of the current analysis, independent of the fact that the resulting variance components scale predictably with this linear adjustment. Third, our definition of statin use based on EHR prescription intervals may misclassify individuals whose prescription history may be incomplete. In these cases, individuals may be classified as untreated despite having received statin therapy that was not captured in the available EHR data. Although this is unlikely to bias genetic analyses, it could affect phenotypic comparison of LDL-C between statin-treated and untreated groups.

Fourth, heritability estimates for individuals on a phenotype modifying treatment are challenging to interpret. We report results for both the unadjusted and statin-adjusted phenotypes because neither is ideal: the unadjusted phenotype is artificially lowered by treatment, while the statin-adjusted value assumes a uniform multiplicative effect that may not be accurate for all individuals. Presenting both provides a range of plausible estimates, but the true heritability in statin-treated individuals likely lies between these two extremes. Additionally, heritability analyses were only performed in EUR-like individuals who additionally self-identify as White and Non-Hispanic limiting the generalizability of heritability results.

Fifth, this study only includes cisgender women and men. Individuals who identify otherwise are not included in this study in part due to low representation in the data set. Sixth, due to large differences in sample size between White and Black individuals, MWU tests comparing racial groups are likely underpowered. Formal interaction testing would require larger, more balanced samples to confirm observed patterns.

Seventh, we did not filter for post-menopausal women on hormone replacement therapy (HRT). Estrogen, specifically estradiol, is well established to convey protective effects for women against CVD [81]. Even though the risk of developing CVDs increases with the decrease in estradiol during menopause partially due to rising LDL-C levels [63], [64], women in the United States are dramatically undertreated with HRT. Practitioners often do not offer HRT to peri-menopausal and menopausal women, and if they do, they are frequently underdosed [82]. Given the low rate of HRT use among peri-menopausal and menopausal women in the United States, we expect that any effect of unaccounted for HRT use on our results is likely to be small, though we cannot rule out some residual influence on LDL-C measurements within this age group.

Eighth, our sex-difference analyses have several limitations. The genome-wide GxSex scan had limited power, and no variant reached genome-wide significance, so we cannot exclude the possibility of further loci with sex-differentiated effects. The interaction scan showed moderate genomic inflation, which may reflect residual population stratification in this mixed-ancestry sample. It might also reflect a residual effect of sex, since covariate-by-sex terms are not included in the interaction model as per standard practice. Our locus-level analysis prioritized loci that were genome-wide significant in the combined non-interaction GWAS for sex-difference testing, and therefore may exclude loci with opposite-signed effects in women and men that are not significant in the pooled analysis. In addition, positional clumping identifies lead variants at distinct loci, but does not evaluate secondary, non-lead signals within the clumping window, which itself does not guarantee statistical independence among the loci tested, potentially affecting multiple-test correction and the interpretation of independent signals. Our phenotype was adjusted for statin use by dividing on-statin LDL-C by a correction factor of 0.7, which assumes that statin effects on LDL-C are the same in women and men. If the effect of statin use differs by sex (e.g., due to differences in statin dosing, adherence, or response by sex), SNP-by-sex interaction test statistics could be biased.

Finally, individuals were stratified by sex to help identify sex-specific genetic effects on LDL-C and sex differences in treatment responses. Stratification may introduce bias, particularly if there are differences in ancestry or genetic variation between women and men. We followed standard practice to minimize this [51].

Future research should build upon these findings by addressing the noted constraints and exploring new avenues. First, expanding the scope of variables to include lifestyle factors (e.g., smoking, diet), detailed anthropometric measures (e.g., waist-to-hip ratios, BMI), and socioeconomic indicators would provide a more comprehensive understanding of LDL-C regulation. Second, including key comorbidities, such as Type 2 Diabetes, which is known to affect LDL-C measurements [83], [84], [85], especially in women [86], [87] may help reveal any sex-specific patterns in comorbidities. Third, broadening participants to include gender-diverse populations is crucial for ensuring that cardiovascular risk screening and treatment guidelines equitably serve these populations. Subsequent analyses should actively include an expanded set of individuals based on the stratified variables and covariates we have considered here to understand population-level health differences. Fourth, more nuanced genetic analyses are warranted. Subsetting individuals by genetic ancestry could reveal variants that are rare or significant in specific ancestral groups. Fifth, it would be beneficial to repeat and extend this analysis in other biobanks to see if results are recapitulated.

In conclusion, we find that while the additive genetic architecture of LDL-C is largely concordant between sexes, the phenotypic distribution, statin treatment rate and therapeutic response, and the residual contributions to LDL-C do differ between women and men, highlighting the need for sex-specific strategies in LDL-C management and cardiovascular disease risk mitigation. Specifically, these strategies should prioritize refined clinical thresholds for women and optimize statin dosing to address the higher residual LDL-C we observed in women. Importantly, the general absence of sex-differences in SNP-based heritability, together with concordant effect sizes at 13 of the 14 distinct lead autosomal loci, underscores that these differences may be driven largely by environmental and clinical management factors—making them amenable to targeted clinical interventions.

## Data and code availability

Individual-level genotype and phenotype data used in this study are available to registered researchers with controlled tier access through the All of Us Researcher Workbench (https://workbench.researchallofus.org/). Access is granted to approved users under the All of Us data use policies.

We performed all QC, phenotypic, and genomic analyses within the All of Us v7 cloud environment using R version 4.4.0 and Python 3.10.12. We used PLINK v1.90b6.22 and v2.0.0-a.6.9LM, GCTA v1.94.4, and REGENIE v2.2.4 and v3.1.4, for genomic analyses. To ensure accessibility for readers with color vision deficiency, we selected color palettes from https://davidmathlogic.com/colorblind.

GWAS and XWAS summary statistics for the combined and sex-stratified analyses are publicly available at https://doi.org/10.5281/zenodo.19333987. R scripts used for post-GWAS analyses will be made available at https://github.com/j-liquori/AoU_LDL_postGWAS upon publication.

## Acknowledgments

First and foremost, we would like to thank the All of Us Research Program participants, without whom this project would not have been possible. We would also like to thank the All of Us Research Program researchers for their recruitment efforts and for building an accessible and easily replicated LDL-C phenotype. Thank you to our colleagues in the Musharoff lab and to Nasa Sinnott-Armstrong for providing constructive discussion and feedback.

## Declaration

The authors declare no competing interests.

