## Supplementary Information for "Phenotypic and genetic sex differences in LDL-C and statin treatment in All of Us"

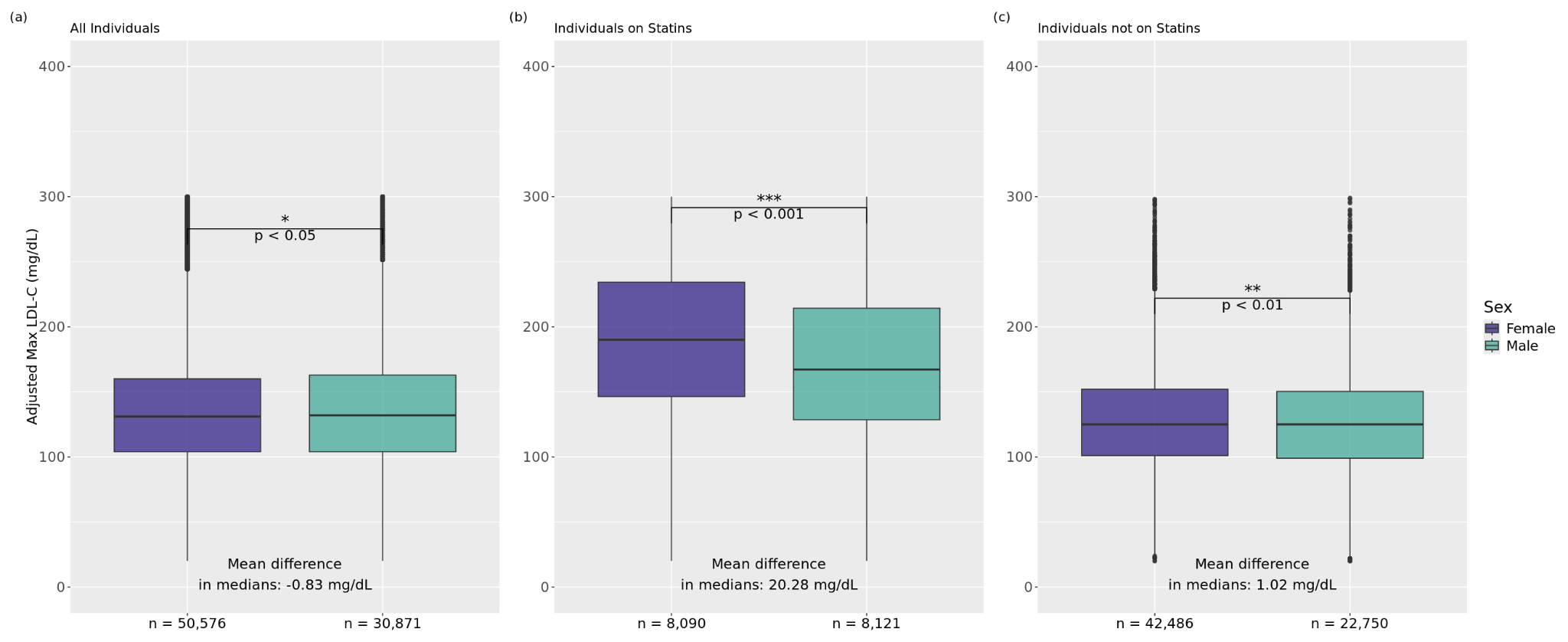
**Figure S1: Statin-adjusted maximum LDL-C levels by sex.** Box plots depicting the distribution of statin-adjusted maximum LDL-C measurements (20–300 mg/dL) in women (purple) and men (green) across three subgroups: (a) all individuals (women: n = 50,576; men: n = 30,871), (b) statin-treated individuals (women: n = 8,090; men: n = 8,121), and (c) untreated individuals (women: n = 42,486; men: n = 22,750). LDL-C values for statin-treated individuals were adjusted using a standard correction factor of 0.7 to approximate untreated LDL-C levels. Boxes represent the interquartile range (IQR), with the horizontal line indicating the median. Whiskers extend to 1.5 x IQR; points beyond this range are plotted individually as outliers. The mean difference in medians as estimated by the Hodges-Lehmann estimator (averaged over 1,000 bootstrap trials) is annotated within each panel. Statistical significance was assessed using the Mann-Whitney U test. In statin-treated individuals, there is a significant difference in medians with women having a mean median increase of 20.28 mg/dL over men. Median differences in non-users and the combined group, while statistically significant, are practically near-null.

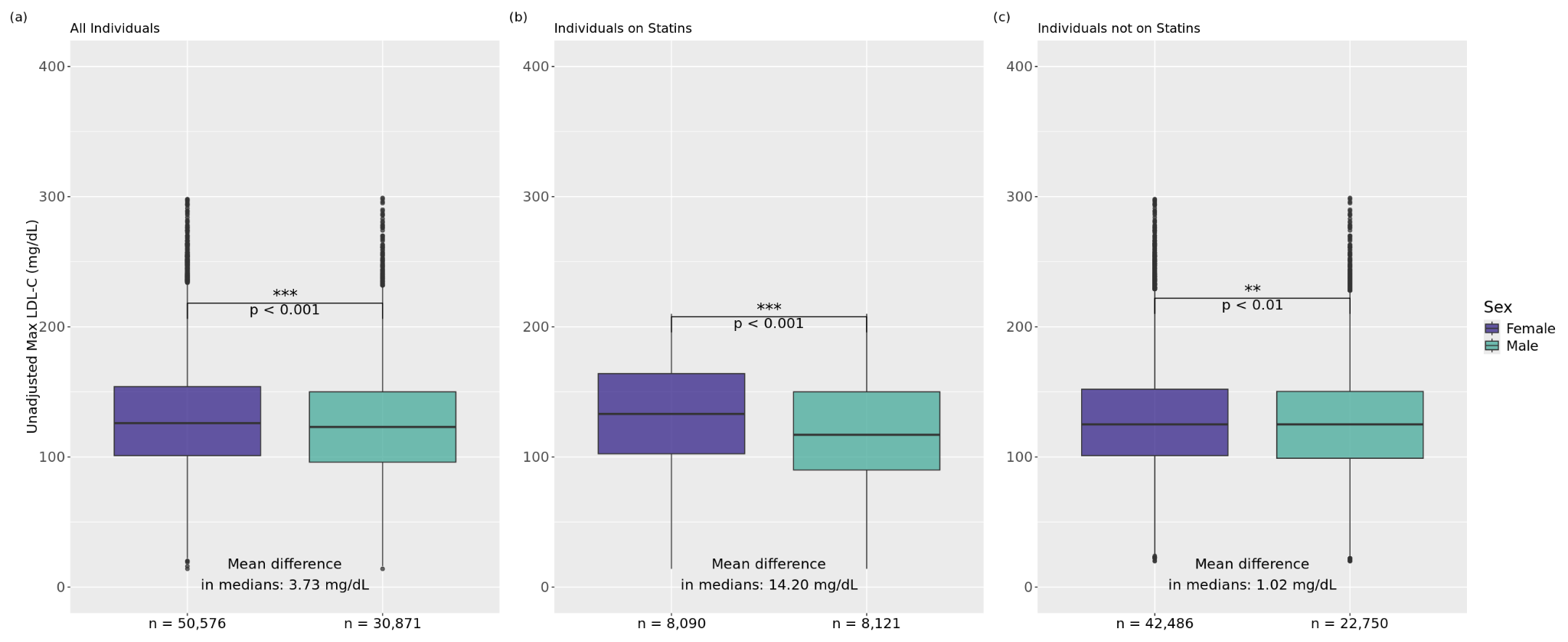
**Figure S2: Unadjusted maximum LDL-C levels by sex.** Box plots depicting the distribution of unadjusted maximum LDL-C measurements (20–300 mg/dL) in women (purple) and men (green) across three subgroups: (a) all individuals (women: n = 50,576; men: n = 30,871), (b) statin-treated individuals (women: n = 8,090; men: n = 8,121), and (c) untreated individuals (women: n = 42,486; men: n = 22,750). Boxes represent the interquartile range (IQR), with the horizontal line indicating the median. Whiskers extend to 1.5 x IQR; points beyond this range are plotted individually as outliers. The mean difference in medians as estimated by the Hodges-Lehmann estimator (averaged over 1,000 bootstrap trials) is annotated within each panel. Statistical significance was assessed using the Mann-Whitney U test. In statin-treated individuals, there is a significant difference in medians with women having a mean median increase of 14.20 mg/dL over men. Median differences in non-users and the combined group, while statistically significant, are practically near-null.

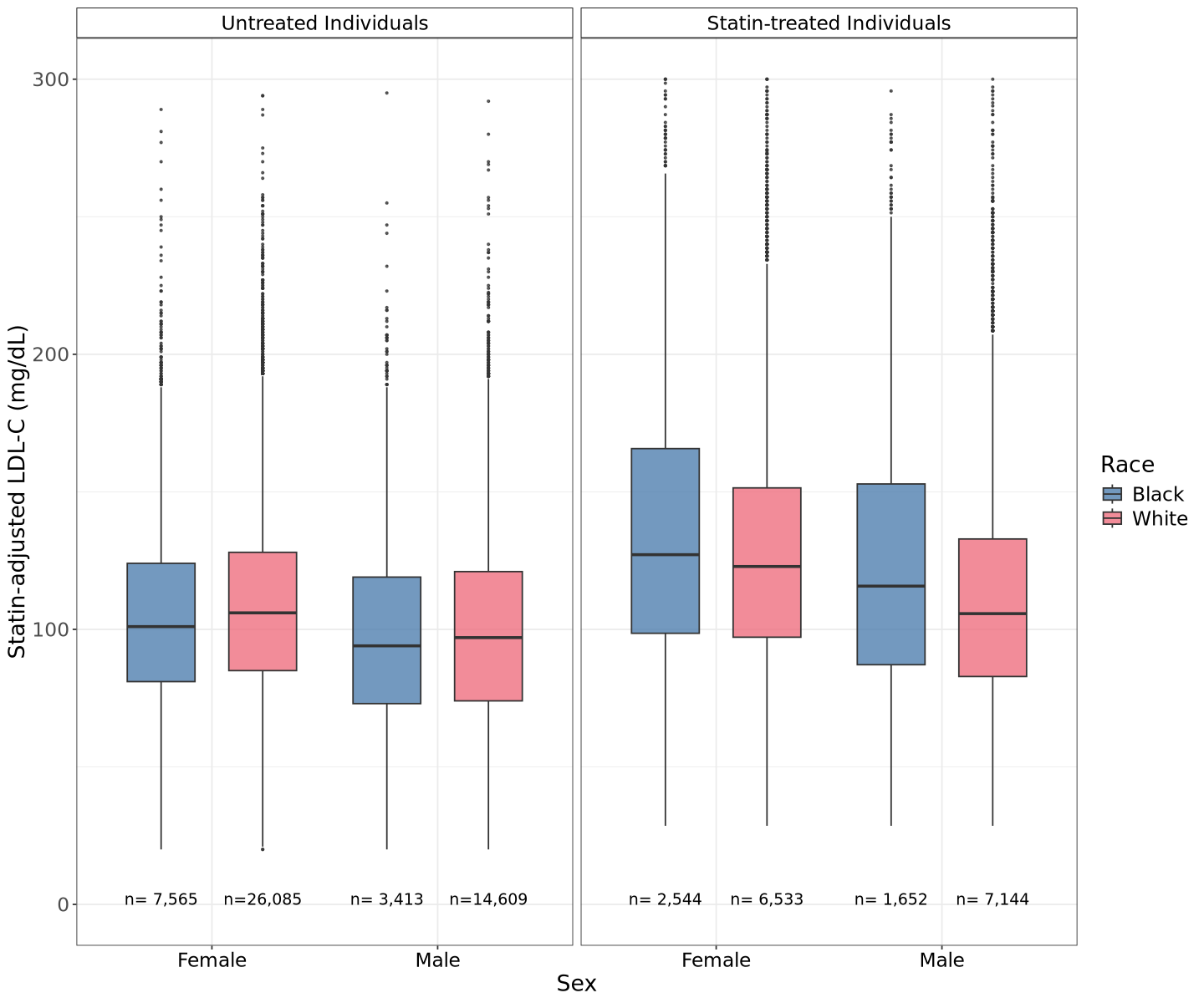
 **Figure S3: Statin-adjusted LDL-C levels by self-reported race, sex, and statin treatment status.** Box plots displaying statin-adjusted LDL-C measurements (mg/dL) for Black (blue) and White (pink) individuals stratified by sex and statin treatment status. Panels show untreated individuals (left) and statin-treated individuals (right). The horizontal line within each box represents the median, boxes represent the interquartile range (IQR), and whiskers extend to 1.5xIQR; points beyond this range are plotted individually as outliers. Sample sizes are indicated below each group. All pairwise comparisons were statistically significant (Mann-Whitney U test, p < 0.001).

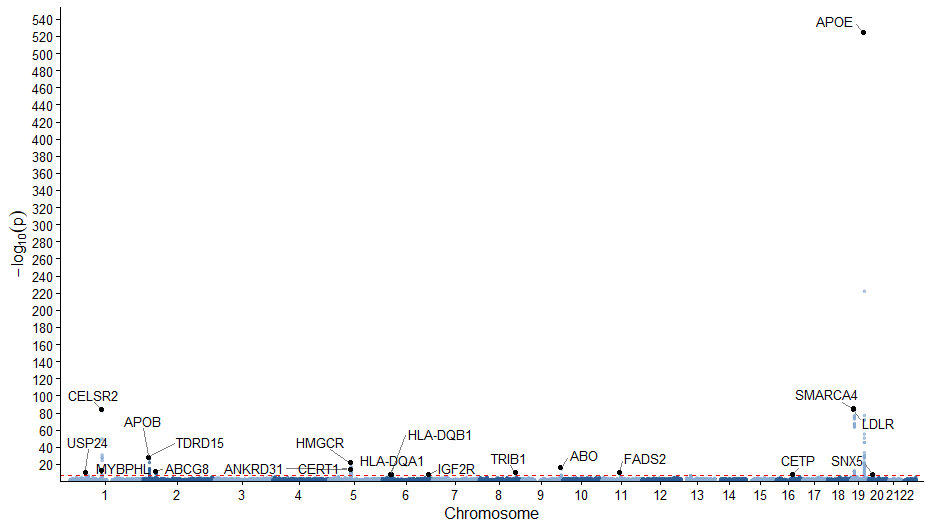

**Figure S4: Manhattan plot of statin-adjusted LDL-C GWAS identifies known trait-associated loci.** Genome-wide significant peaks replicate known LDL-C associated loci; the dashed red line indicates the p-value cutoff based on Bonferroni adjustment, and all results are derived from the combined analysis group.

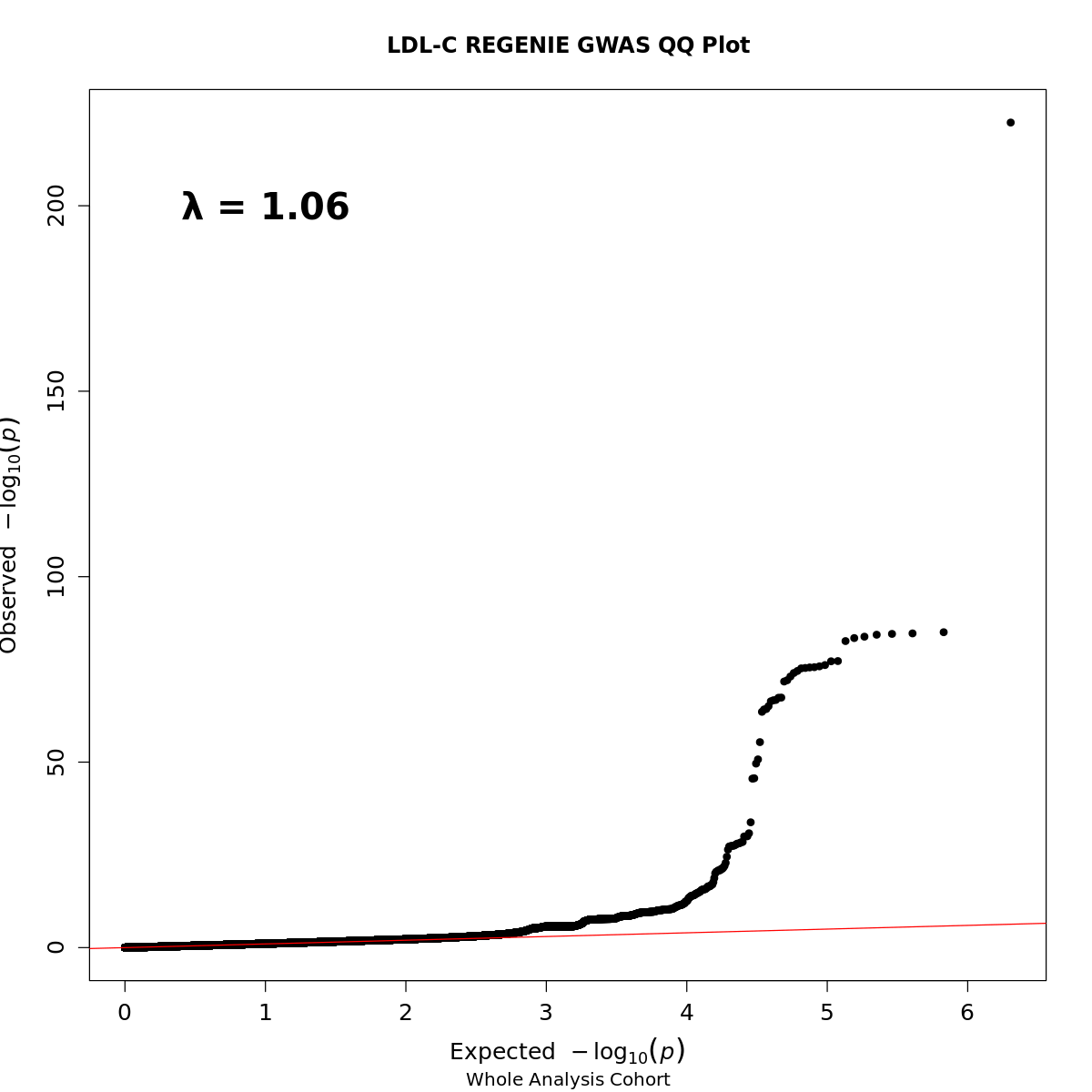

**Figure S5: QQ Plot of the LDL-C GWAS on the combined analysis group shows associations with minimal inflation (λ = 1.06).** Observed versus expected -log_10_(*p*) values under the null hypothesis are plotted for the combined analysis group GWAS; the red line represents the null expectation (y=x). Most points closely follow the diagonal indicating well-calibrated test statistics. The upward deviation in the upper tail is consistent with true signals rather than systematic bias. The genomic inflation factor is low (λ = 1.06) indicating any residual inflation is minimal.

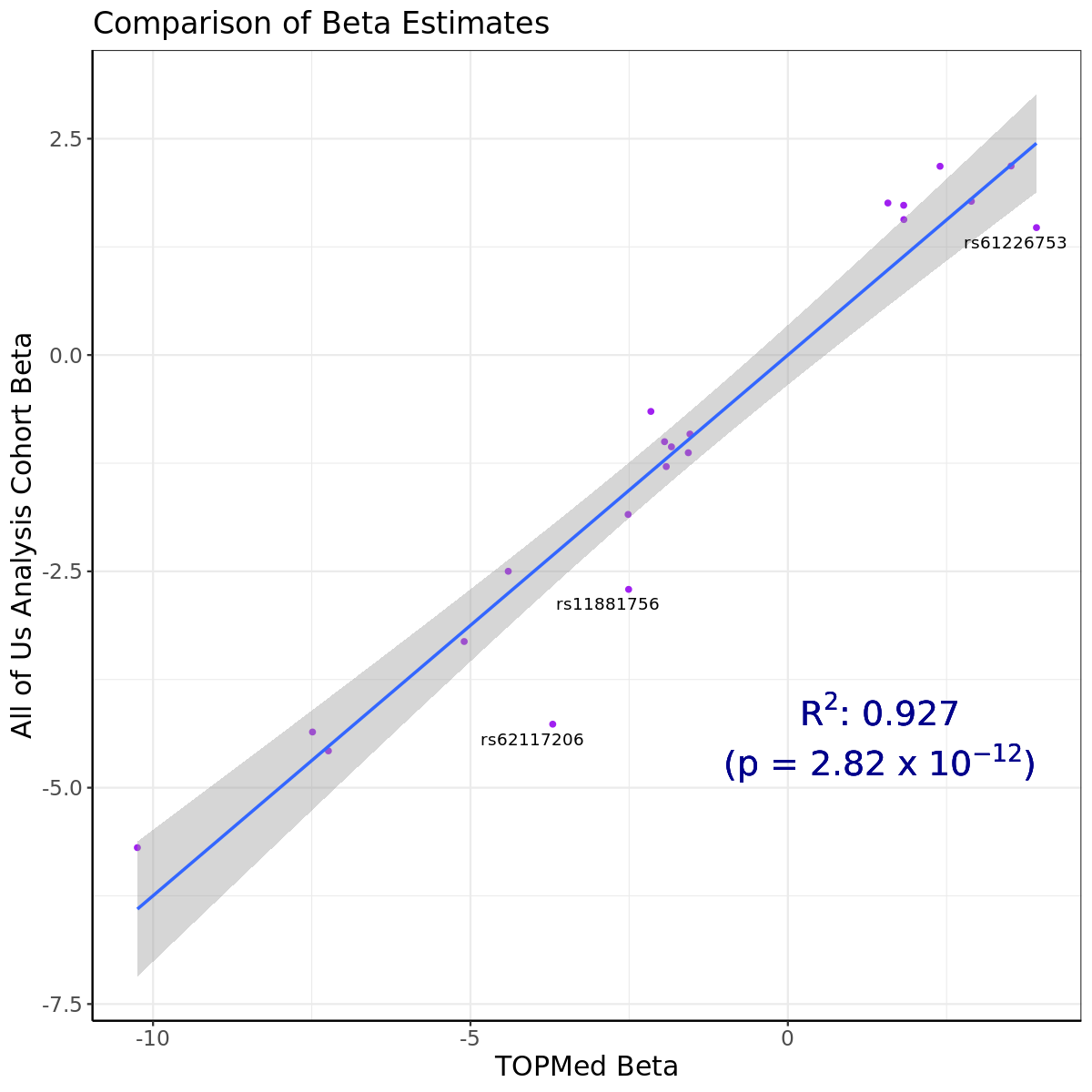

**Figure S6: Combined analysis group LDL-C GWAS effect size estimates show a strong positive association with previously reported effect size estimates.** This scatterplot demonstrates a strong positive correlation between the effect size estimates from the combined analysis group GWAS (y-axis), using untransformed LDL-C, and previously reported effect size estimates from the TOPMed dataset (x-axis) [[22]](https://www.zotero.org/google-docs/?oh2jYa), which uses a rank inverse normalized transformed phenotype. The R² value of 0.927 indicates a high degree of agreement with a highly significant *p*-value (*p* = 2.8e-12) supporting the robustness of findings in the combined analysis group. This is a replication of the comparison found in the *All of Us* Research Program’s exploration of LDL-C [[21]](https://www.zotero.org/google-docs/?C2WAsj), which we repeated to ensure the consistency of our results.

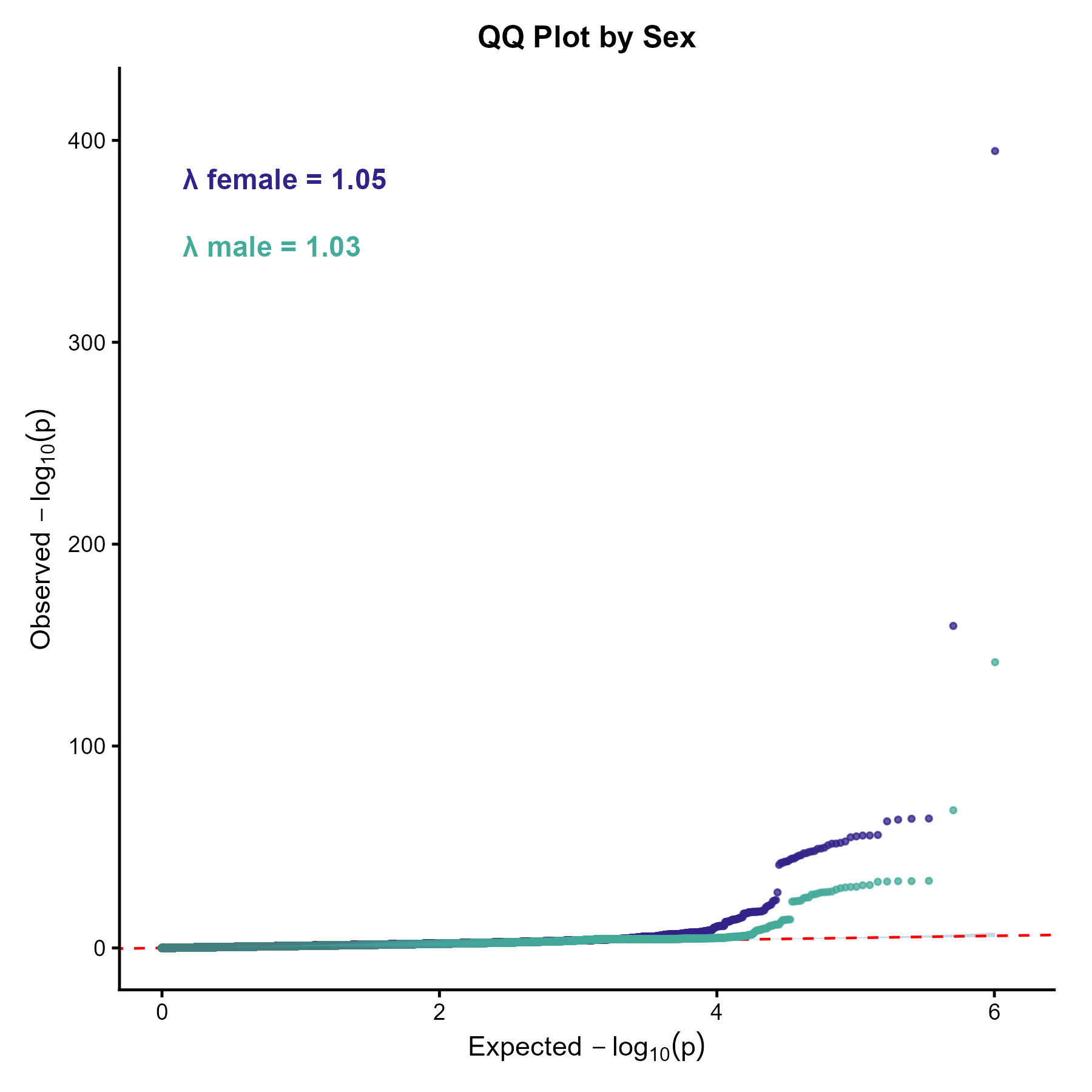

**Figure S7: QQ plots of the sex-stratified GWASs of LDL-C.** Observed versus expected -log_10_(*p*) values under the null hypothesis are plotted for women (purple) and men (green); the red dashed line represents the null expectation (y=x). Most points closely follow the diagonal indicating well-calibrated test statistics. The slight upward deviation in the upper tail is consistent with true signals rather than systematic bias. Genomic inflation factors are low (female λ = 1.05, male λ = 1.03) indicating any residual inflation is minimal.

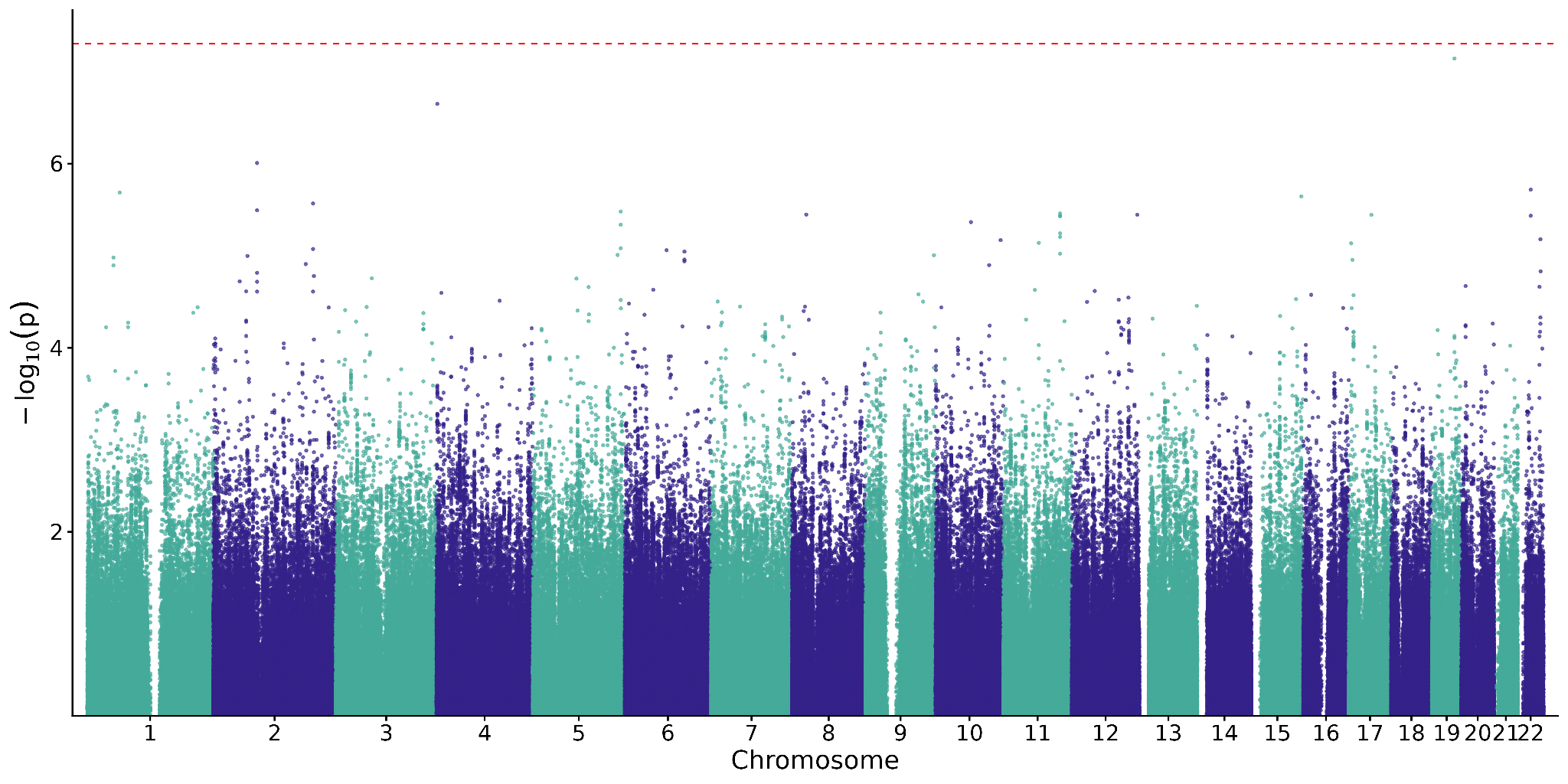

**Figure S8: Manhattan plot of GxSex GWAS of statin-adjusted LDL-C.** The dashed red line indicates the p-value cutoff based on Bonferroni adjustment, and all results are derived from the sex-pooled group. No variants show a statistically significant SNP-by-sex effect after Bonferroni correction.

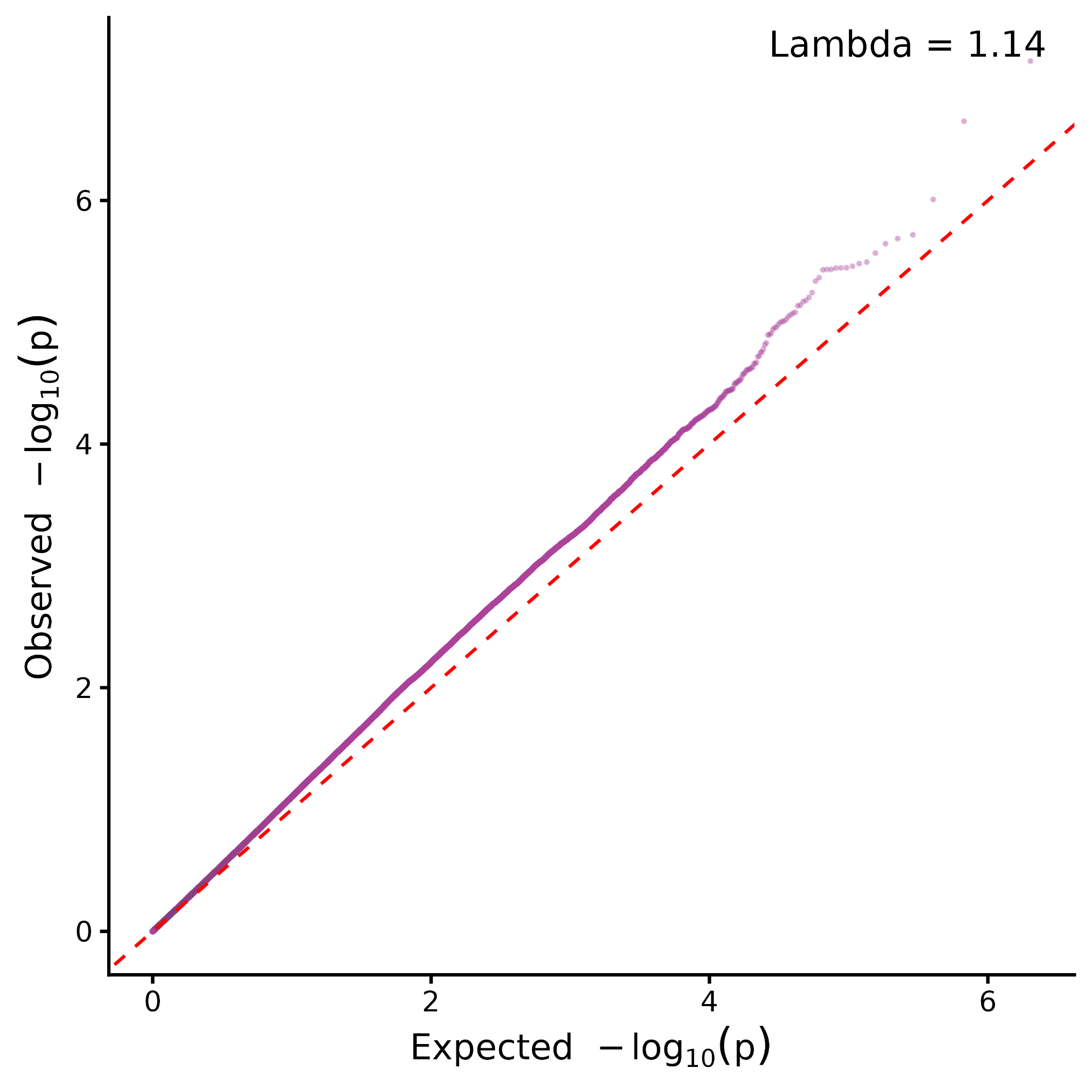

**Figure S9: QQ plot of the GxSex GWAS of statin-adjusted LDL-C.** Observed versus expected -log_10_(*p*) values under the null hypothesis are plotted for the sex-pooled group; the red dashed line represents the null expectation (y=x). Most points closely follow the diagonal indicating well-calibrated test statistics. Genomic inflation factor is moderate (λ = 1.14) indicating mild residual inflation.

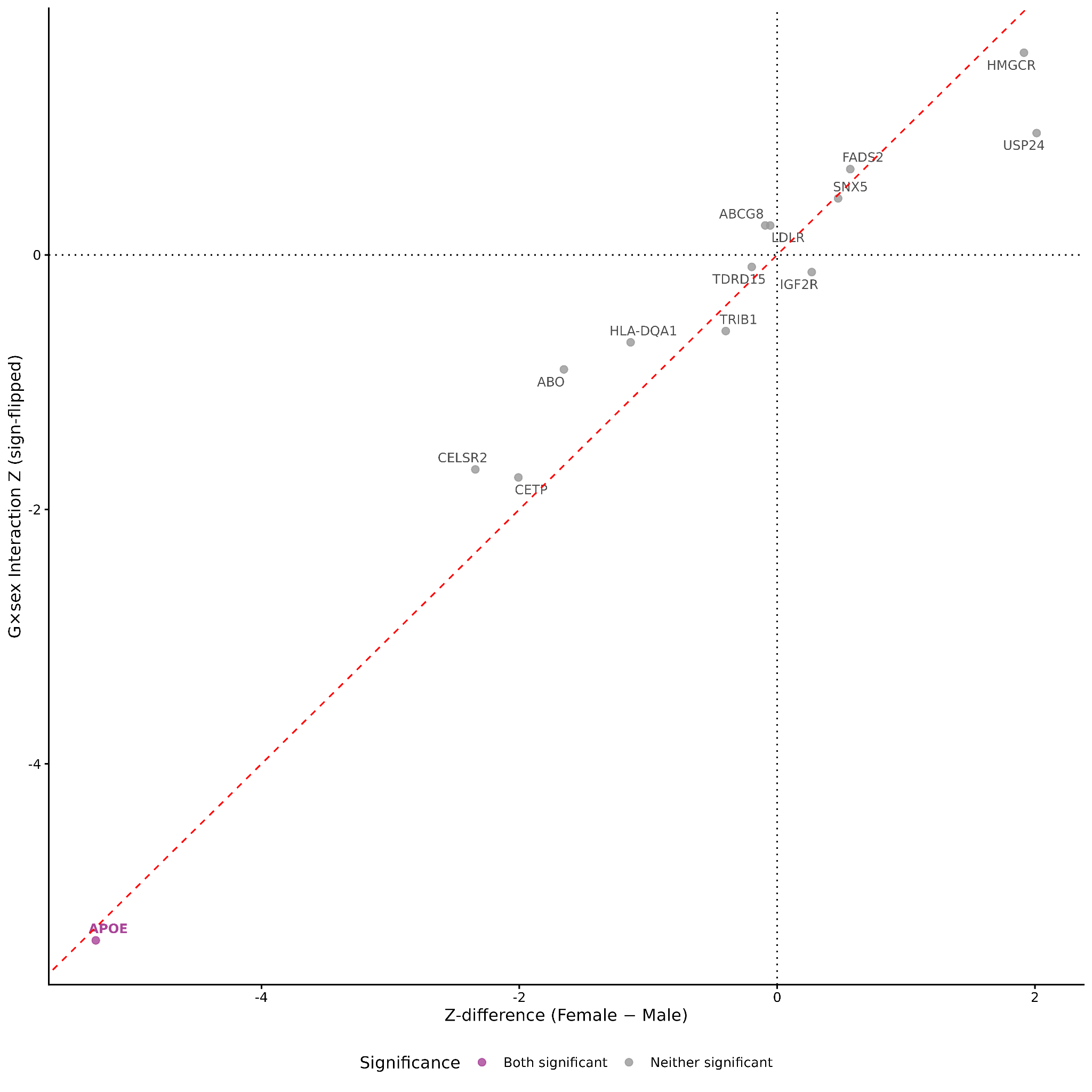

**Figure S10: Concordance between sex-stratified GWAS and GxSex interaction results at 14 lead LDL-C loci.** Scatterplot comparing sex-difference Z-scores from sex-stratified GWAS (x-axis, female minus male) with Z-scores from pooled GxSex interaction tests (y-axis, sign-flipped) across 14 lead loci. The red dashed line indicates y = x, and the dotted horizontal and vertical lines indicate zero. Points are colored according to statistical significance in the sex-differences and GxSex interaction analyses, with gene labels identifying the 14 lead loci. *APOE/APOC1* was the only locus showing evidence of sex differences in both analyses and remained significant after Bonferroni correction across the 14 lead loci, although it did not meet the genome-wide Bonferroni threshold in the formal GxSex analysis.

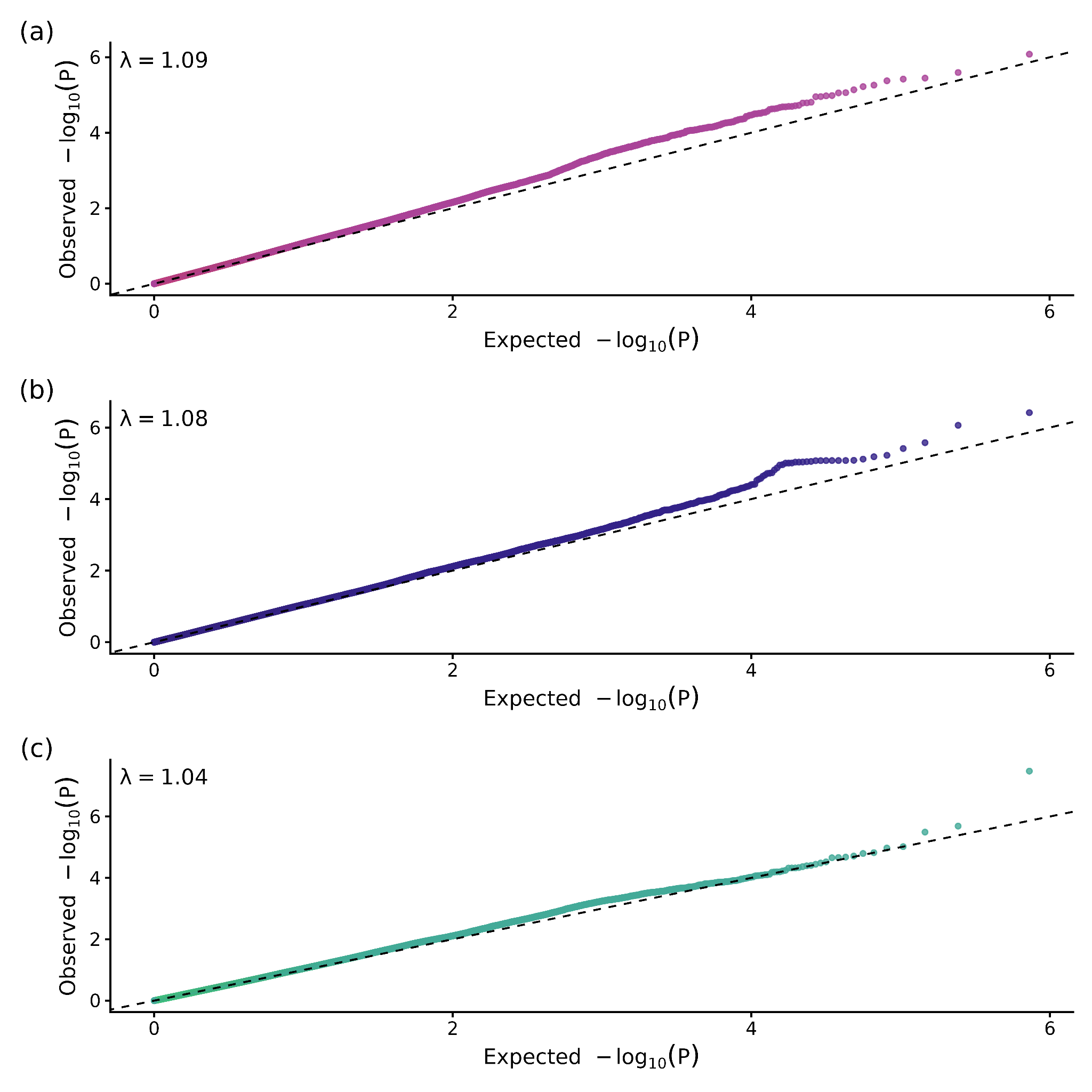

**Figure S11: QQ plots of XWASs of LDL-C.** Observed versus expected -log_10_(*p*) values under the null hypothesis are plotted for the (a) Combined analysis group (pink) analysis, (b) women (purple) only and, (c) men only (green); the dashed line represents the null expectation (y=x). Most points closely follow the diagonal indicating well-calibrated test statistics. Genomic inflation factors are acceptable (λ = 1.09, 1.08, and 1.04, respectively) indicating any residual inflation is minimal.

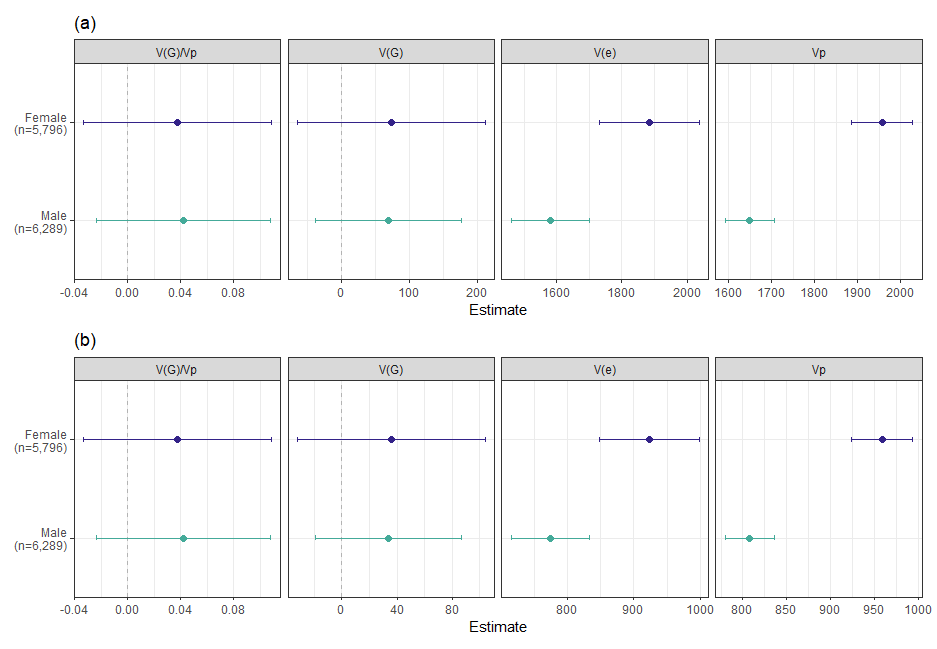
**Figure S12: SNP-based heritability and variance components of LDL-C by sex (statin-treated individuals).** Forest plot showing the SNP-based heritability (leftmost panel V(G)/Vp), additive genetic variance (second panel, V(G)), residual variance (third panel, V(e)), and phenotypic variance (rightmost panel, Vp) estimates for statin-treated women (purple) and men (green) for (a) Statin-adjusted LDL-C and (b) Unadjusted LDL-C. Heritability and additive genetic variance do not differ significantly between women and men as confirmed by a z-score difference test. However, V(e) was significantly (Z_diff_ = 3.07, p-value = 0.002) higher in women (adjusted LDL-C V(e) =1884, unadjusted LDL-C V(e) = 923) than in men (adjusted LDL-C V(e) = 1580, unadjusted LDL-C V(e) = 774). Additionally, Vp differs between sexes across all groups (all p ≤ 0.0003), being higher in women (adjusted LDL-C Vp = 1957, unadjusted LDL-C Vp = 959) than in men (adjusted LDL-C Vp = 1649, unadjusted LDL-C Vp = 808).

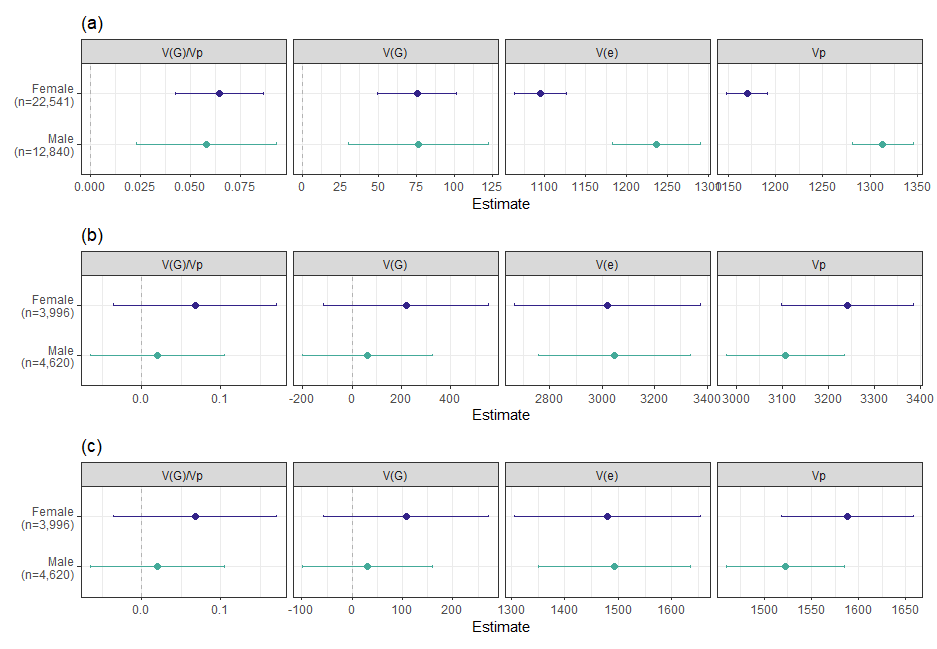

**Figure S13: SNP-based heritability and variance components by sex for maximum LDL-C (20–300 mg/dL).** Forest plot showing the SNP-based heritability (leftmost panel V(G)/Vp), additive genetic variance (second panel, V(G)), residual variance (third panel, V(e)), and phenotypic variance (rightmost panel, Vp) estimates for statin-treated women (purple) and men (green) for (a) Unadjusted LDL-C in untreated individuals, (b) Statin-adjusted LDL-C in statin-treated individuals, and (c) Unadjusted LDL-C in statin-treated individuals. Heritability and additive genetic variance do not differ significantly between women and men as confirmed by a z-score difference test. However, among untreated individuals, unadjusted LDL-C V(e) was significantly (Z_diff_ = -4.45, p-value = 8.4e-6) lower in women (unadjusted LDL-C V(e) = 1095) than in men (unadjusted LDL-C V(e) = 1237). In statin-treated individuals, V(e) did not differ significantly between sexes. Additionally, Vp differed significantly between sexes among untreated individuals (Z_diff_ = -7.21, p-value = 5.5e-13), with lower values in women (Vp = 1170) than in men (Vp = 1313). In statin-treated individuals, Vp did not differ significantly between sexes.

| **Characteristic** | **Number of individuals** |
| --- | --- |
| **Predicted Genetic Ancestry** |  |
| African | 16,024 |
| American | 9,652 |
| East Asian | 1,616 |
| European | 51,387 |
| Middle Eastern | 214 |
| South Asian | 852 |
| Other | 6,955 |
| **Self-Reported Ethnic Identity** | |
| Hispanic or Latino | 11,881 |
| Not Hispanic or Latino | 72,509 |
| None Selected | 2,310 |
| **Self-Reported Racial Identity** | |
| Asian | 2,421 |
| Black or African American | 15,174 |
| Middle Eastern or North African | 506 |
| More Than One Population | 1,306 |
| Native Hawaiian or Other Pacific Islander | 54 |
| White | 54,371 |
| None Selected | 12,868 |

**Table S1. Sample characteristics.** Table with the number of individuals within each self-identified race and ethnicity group and AoU predicted ancestries.

| **Group (Unadjusted LDL-C)** | **Female Median (n)** | **Male Median (n)** | **KL Divergence** | **HL Estimate** | **95% CI** | **MWU p-value** |
| --- | --- | --- | --- | --- | --- | --- |
| All Individuals | 101.00 mg/dL (n = 54,189) | 89.00 mg/dL (n = 32,511) | 0.05042 | 11.42 mg/dL | 11.00 to 12.00 mg/dL | 2.2e-16 |
| Statin-treatedIndividuals | 87.00 mg/dL (n = 10,977) | 75.00 mg/dL (n = 10,253) | 0.07703 | 11.62 mg/dL | 11.00 to 12.00 mg/dL | 1.014e-167 |
| Untreated Individuals | 105.00 mg/dL (n = 43,212) | 97.00 mg/dL (n = 22,258) | 0.02582 | 8.09 mg/dL | 8.00 to 9.00 mg/dL | 2.816e-182 |

**Table S2. Summary of statistical tests of unadjusted LDL-C.** This table shows the values obtained from performing a Kullback-Leibler divergence test, the Mann-Whitney U test, and the Hodges-Lehman estimate on unadjusted LDL-C measurements from women and men. The table also includes the 95% confidence interval for the Hodges-Lehmann estimator.

| **Group (Adjusted LDL-C)** | **Female Median (n)** | **Male Median (n)** | **KL Divergence** | **HL Estimate** | **95% CI** | **MWU p-value** |
| --- | --- | --- | --- | --- | --- | --- |
| All Individuals | 108.00 mg/dL (n = 54,189) | 100.00 mg/dL (n = 32,511) | 0.02571 | 8.18 mg/dL | 8.00 to 9.00 mg/dL | 3.333e-227 |
| Statin-treatedIndividuals | 124.29 mg/dL (n = 10,977) | 107.14 mg/dL (n = 10,253) | 0.07703 | 16.60 mg/dL | 15.71 to 17.14 mg/dL | 1.014e-167 |
| Untreated Individuals | 105.00 mg/dL (n = 43,212) | 97.00 mg/dL (n = 22,258) | 0.02582 | 8.09 mg/dL | 8.00 to 9.00 mg/dL | 2.816e-182 |

**Table S3. Summary of statistical tests of adjusted LDL-C.** This table shows the values obtained from performing a Kullback-Leibler divergence test, the Mann-Whitney U test, and the Hodges-Lehman estimate on statin-adjusted LDL-C measurements from women and men. The table also includes the 95% confidence interval for the Hodges-Lehmann estimator.

| **Subgroup** | **Female Median mg/dL (n)** | **Male Median mg/dL (n)** | **KL Div.** | **HL Estimate, mg/dL (95% CI)** | **MWU p-value** |
| --- | --- | --- | --- | --- | --- |
| **Statin-adjusted maximum LDL-C (20–300 mg/dL)** | | | | | |
| All Individuals | 131.00 (n=50,576) | 132.00 (n=30,871) | 0.004 | -0.83 (-1.43-0.00) | 1.277e-02 |
| Statin-treated Individuals | 190.00 (n=8,090) | 167.14 (n=8,121) | 0.055 | 20.28 (18.57-21.43) | 1.691e-97 |
| Untreated Individuals | 125.00 (n=42,486) | 125.00 (n=22,750) | 0.004 | 1.02 (0.00-2.00) | 1.089e-03 |
| **Unadjusted maximum LDL-C (20-300 mg/dL)** | | | | | |
| All Individuals | 126.00 (n=50,576) | 123.00 (n=30,871) | 0.008 | 3.73 (3.00-4.00) | 1.930e-38 |
| Statin-Treated Individuals | 133.00 (n=8,090) | 117.00 (n=8,121) | 0.055 | 14.20 (13.00-15.00) | 1.691e-97 |
| Untreated Individuals | 125.00 (n=42,486) | 125.00 (n=22,750) | 0.004 | 1.02 (0.00- 2.00) | 1.089e-03 |

**Table S4. Summary of statistical tests of maximum LDL-C (20–300 mg/dL).** This table shows the values obtained from performing a Kullback-Leibler divergence test, the Mann-Whitney U test, and the Hodges-Lehman estimate on statin-adjusted and unadjusted maximum LDL-C measurements from women and men. The table also includes the 95% confidence interval for the Hodges-Lehmann estimator.

| **Phenotype** | **Subgroup** | **Female Median mg/dL (n)** | **Male Median mg/dL (n)** | **KL Div.** | **HL Estimate, mg/dL (95% CI)** | **MWU p-value** |
| --- | --- | --- | --- | --- | --- | --- |
| **Statin-adjusted LDL-C (< 1,000 mg/dL)** | | | | | | |
| Most Recent | All | 108.00 (n=54,578) | 100.00 (n=32,826) | 0.408 | 8.50 (8.00-9.00) | 2.799e-239 |
| Maximum | All | 132.00 (n=51,212) | 133.00 (n=31,182) | 0.780 | -0.54 (-1.00-0.00) | 8.846e-02 |
| Most Recent | Untreated | 105.00 (n=43,312) | 97.00 (n=22,370) | 0.039 | 8.16 (8.00-9.00) | 2.462e-186 |
| Maximum | Untreated | 125.00 (n=42,531) | 125.00 (n=22,779) | 1.921 | 1.02 (0.00-2.00) | 6.953e-04 |
| Most Recent | Statin-treated | 124.29 (n=11,266) | 107.14 (n=10,456) | 0.215 | 17.38 (15.71-18.57) | 9.018e-181 |
| Maximum | Statin-treated | 197.14 (n=8,681) | 170.00 (n=8,403) | 0.037 | 23.99 (21.43-25.71) | 1.944e-120 |
| **Statin-adjusted LDL-C (10-500 mg/dL)** | | | | | | |
| Most Recent | All | 108.00 (n=54,575) | 100.00 (n=32,826) | 0.064 | 8.51 (8.00-9.00) | 4.184e-239 |
| Maximum | All | 132.00 (n=51,189) | 133.00 (n=31,177) | 0.004 | -0.58 (-1.00-0.00) | 7.606e-02 |
| Most Recent | Untreated | 105.00 (n=43,312) | 97.00 (n=22,370) | 0.040 | 8.16 (8.00-9.00) | 2.462e-186 |
| Maximum | Untreated | 125.00 (n=42,527) | 125.00 (n=22,779) | 0.003 | 1.02 (0.00-2.00) | 7.467e-04 |
| Most Recent | Statin-treated | 124.29 (n=11,263) | 107.14 (n=10,456) | 0.007 | 17.35 (15.71-18.57) | 2.024e-180 |
| Maximum | Statin-treated | 197.14 (n=8,662) | 170.00 (n=8,398) | 0.080 | 23.86 (21.43-25.71) | 4.204e-119 |
| **Statin-adjusted LDL-C (20-300 mg/dL)** | | | | | | |
| Most Recent | All | 108.00 (n=54,189) | 100.00 (n=32,511) | 0.026 | 8.18 (8.00-9.00) | 3.333e-227 |
| Maximum | All | 131.00 (n=50,576) | 132.00 (n=30,871) | 0.003 | -0.83 (-1.43-0.00) | 1.277e-02 |
| Most Recent | Untreated | 105.00 (n=43,212) | 97.00 (n=22,258) | 0.026 | 8.09 (8.00-9.00) | 2.816e-182 |
| Maximum | Untreated | 125.00 (n=42,486) | 125.00 (n=22,750) | 0.004 | 1.02 (0.00-2.00) | 1.089e-03 |
| Most Recent | Statin-treated | 124.29 (n=10,977) | 107.14 (n=10,253) | 0.077 | 16.60 (15.71-17.14) | 1.014e-167 |
| Maximum | Statin-treated | 190.00 (n=8,090) | 167.14 (n=8,121) | 0.055 | 20.28 (18.57-21.43) | 1.691e-97 |

**Table S5: Sensitivity analysis of sex differences in statin-adjusted LDL-C across most recent and maximum measurements.** Median differences, MWU p-values, and KL divergence for women and men across filtration ranges, phenotypes, and statin usage subgroups.

| **Phenotype** | **Subgroup** | **Female Median mg/dL (n)** | **Male Median mg/dL (n)** | **KL Div.** | **HL Estimate, mg/dL (95% CI)** | **MWU p-value** |
| --- | --- | --- | --- | --- | --- | --- |
| **Unadjusted LDL-C (< 1,000 mg/dL)** | | | | | | |
| Most Recent | All | 101.00 (n=54,578) | 89.00 (n=32,826) | 0.023 | 11.75 (11.00-12.00) | 0e+00 |
| Maximum | All | 127.00 (n=51,212) | 123.00 (n=31,182) | 1.694 | 3.92 (3.00-4.00) | 1.703e-41 |
| Most Recent | Untreated | 105.00 (n=43,312) | 97.00 (n=22,370) | 0.039 | 8.16 (8.00-9.00) | 2.462e-186 |
| Maximum | Untreated | 125.00 (n=42,531) | 125.00 (n=22,779) | 1.921 | 1.02 (0.00-2.00) | 6.953e-04 |
| Most Recent | Statin-treated | 87.00 (n=11,266) | 75.00 (n=10,456) | 0.215 | 12.17 (11.00-13.00) | 9.018e-181 |
| Maximum | Statin-treated | 138.00 (n=8,681) | 119.00 (n=8,403) | 0.037 | 16.79 (15.00-18.00) | 1.944e-120 |
| **Unadjusted LDL-C (10-500 mg/dL)** | | | | | | |
| Most Recent | All | 101.00 (n=54,575) | 89.00 (n=32,826) | 0.022 | 11.77 (11.00-12.00) | 0e+00 |
| Maximum | All | 127.00 (n=51,189) | 123.00 (n=31,177) | 0.009 | 3.88 (3.00-4.00) | 4.010e-41 |
| Most Recent | Untreated | 105.00 (n=43,312) | 97.00 (n=22,370) | 0.039 | 8.16 (8.00-9.00) | 2.462e-186 |
| Maximum | Untreated | 125.00 (n=42,527) | 125.00 (n=22,779) | 0.003 | 1.02 (0.00-2.00) | 7.467e-04 |
| Most Recent | Statin-treated | 87.00 (n=11,263) | 75.00 (n=10,456) | 0.007 | 12.14 (11.00-13.00) | 2.024e-180 |
| Maximum | Statin-treated | 138.00 (n=8,662) | 119.00 (n=8,398) | 0.080 | 16.71 (15.00-18.00) | 4.204e-119 |
| **Unadjusted LDL-C (20-300 mg/dL)** | | | | | | |
| Most Recent | All | 101.00 (n=54,189) | 89.00 (n=32,511) | 0.050 | 11.42 (11.00-12.00) | 0e+00 |
| Maximum | All | 126.00 (n=50,576) | 123.00 (n=30,871) | 0.008 | 3.73 (3.00-4.00) | 1.930e-38 |
| Most Recent | Untreated | 105.00 (n=43,212) | 97.00 (n=22,258) | 0.026 | 8.09 (8.00-9.00) | 2.816e-182 |
| Maximum | Untreated | 125.00 (n=42,486) | 125.00 (n=22,750) | 0.004 | 1.02 (0.00-2.00) | 1.089e-03 |
| Most Recent | Statin-treated | 87.00 (n=10,977) | 75.00 (n=10,253) | 0.077 | 11.62 (11.00-12.00) | 1.014e-167 |
| Maximum | Statin-treated | 133.00 (n=8,090) | 117.00 (n=8,121) | 0.055 | 14.20 (13.00-15.00) | 1.691e-97 |

**Table S6: Sensitivity analysis of sex differences in unadjusted LDL-C across most recent and maximum measurements.** Median differences, MWU p-values, and KL divergence for women and men across filtration ranges, phenotypes, and statin usage subgroups.

| **Age Group** | **Female Median mg/dL (treated n)** | **Female Total n** | **Female Treatment Rate** | **Male Median mg/dL (treated n)** | **Male Total n** | **Male Treatment Rate** |
| --- | --- | --- | --- | --- | --- | --- |
| 21-30 | 119.30 (44) | 4,469 | 0.98% | 127.10 (29) | 1,794 | 1.62% |
| 31-40 | 137.10 (233) | 7,070 | 3.30% | 127.1 (199) | 3,178 | 6.26% |
| 41-50 | 130.0 (881) | 8,775 | 10.04% | 124.30 (712) | 4,027 | 17.68% |
| 51-60 | 131.40 (2,579) | 12,267 | 21.02% | 115.70 (2,068) | 6,702 | 30.86% |
| 61-70 | 122.90 (4,094) | 13,342 | 30.69% | 107.10 (3,652) | 9,176 | 39.80% |
| 71-80 | 118.60 (3,146) | 8,266 | 38.06% | 100.00 (3,593) | 7,634 | 47.07% |

**Table S7. Median LDL-C and statin treatment rates of statin-treated individuals by sex and age group.** This table shows the statin treatment rate and higher median LDL-C measurements of individuals treated with statins grouped by sex and age group. Across all age strata, men are treated with statins at a higher rate than women despite women typically having higher median LDL-C measurements. Statin treatment rate is calculated as the number of statin-treated individuals within each sex and age group divided by the total number of individuals in that sex and age group.

| **Age Group** | **Female Median mg/dL (untreated n)** | **Male Median mg/dL (untreated n)** |
| --- | --- | --- |
| 21-30 | 95.00 (4,425) | 98.00 (1,765) |
| 31-40 | 100.00 (6,837) | 105.70 (2,979) |
| 41-50 | 106.00 (7,894) | 106.00 (3,315) |
| 51-60 | 112.00 (9,688) | 100.0 (4,634) |
| 61-70 | 109.00 (9,248) | 92.00 (5,524) |
| 71-80 | 100.00 (5,120) | 84.00 (4,041) |

**Table S8. Median LDL-C of untreated individuals by sex and age group.** This table shows the higher median LDL-C measurements of individuals not treated with statins grouped by sex and age group.

| **Stratification** | **Group 1: Median mg/dL (n)** | **Group 2: Median mg/dL (n)** | **HL mg/dL (95% CI)** | **MWU p-value** |
| --- | --- | --- | --- | --- |
| **Black versus White** | | | | |
| Female, Untreated | 101.00 (7,565) | 106.00 (26,085) | -4.63 (-5.00- -4.00) | < 2.2e-16 |
| Female, Statin-treated | 127.14 (2,544) | 122.86 (6,533) | 6.05 (4.29-8.57) | 2.22e-8 |
| Male, Untreated | 94.00 (3,413) | 97.00 (14,609) | -1.73 (-3.00-0.00) | 8.41e-3 |
| Male, Statin-treated | 115.71 (1,652) | 105.71 (7,144) | 9.45 (7.14-11.43) | 2.74e-15 |
| **Female versus Male** | | | | |
| Black, Untreated | 101.00 (7,565) | 94.00 (3,413) | 6.59 (5.00-8.00) | < 2.2e-16 |
| Black, Statin-treated | 127.14 (2,544) | 115.71 (1,652) | 12.79 (10.00-15.71) | < 2.2e-16 |
| White, Untreated | 106.00 (26,085) | 97.00 (14,609) | 9.40 (9.00-10.00) | < 2.2e-16 |
| White, Statin-treated | 122.86 (6,533) | 105.71 (7,144) | 16.29 (14.29-17.14) | < 2.2e-16 |
| **Untreated versus Statin-treated** | | | | |
| Black, Female | 101.00 (7,565) | 127.14 (2,544) | -27.81(-30.00- -25.71) | < 2.2e-16 |
| Black, Male | 94.00 (3,413) | 115.71 (1,652) | -21.63 (-24.43- -19.00) | < 2.2e-16 |
| White, Female | 106.00 (26,085) | 122.86 (6,533) | -17.22 (-18.29- -16.14) | < 2.2e-16 |
| White, Male | 97.00 (14,609) | 105.71 (7,144) | -10.27 (-11.29- -9.29) | < 2.2e-16 |

**Table S9: Pairwise comparisons of statin-adjusted LDL-C by race, sex, and statin treatment status.** Results of Mann-Whitney U (MWU) tests for 12 pairwise comparisons of statin-adjusted LDL-C measurements (mg/dL) across three comparison types: self-reported race (Black vs White), sex (Female vs Male), and statin treatment status (treated vs untreated), each stratified by the remaining two variables. Group 1 and Group 2 columns report the subgroup, sample size (n), and median LDL-C. The Hodges-Lehmann (HL) estimate represents the mean difference in medians between the subgroups averaged over 1,000 bootstrap trials, with the 95% confidence interval. A negative HL estimate indicates Group 1 had a lower median LDL-C than Group 2. P-values are from the MWU tests; values below machine epsilon (2.2e-16) are reported as < 2.2e-16.

| **Model** | **Total R²** | **Partial R² for method** | **Incremental R²** |
| --- | --- | --- | --- |
| Reduced (no method) | 0.0293 | - | - |
| Full (including method) | 0.0326 | 0.0035 | 0.0033 |

**Table S10. Variance in statin-adjusted LDL-C explained by measurement method.** Models include all GWAS covariates. The reduced model omits measurement method; the full model includes method as a categorical variable. Partial R^2^ = (RSS_reduced_ - RSS_full_)/RSS_reduced_. Incremental R^2^ = RSS_full_ - RSS_reduced_.

| **LDL-C Measurement Method** | **Female %** | **Male %** |
| --- | --- | --- |
| Calculated | 66.3% | 67.5% |
| Direct | 4.7% | 6.5% |
| Electrophoresis | 3.5% | 3.4% |
| Ultracentrifugation | 0.6% | 0.3% |
| Unspecified | 24.8% | 22.3% |

**Table S11. Proportion of LDL-C measurement methods by Sex.** Values represent the fraction of individuals within each sex assigned to each measurement method category.

| **Chr** | **Position** | **Variant** | **Gene** | **Combined** | | | **Female** | | | **Male** | | | **Sex Difference** | | |
| --- | --- | --- | --- | --- | --- | --- | --- | --- | --- | --- | --- | --- | --- | --- | --- |
|  |  |  |  | **Beta** | **SE** | **P** | **Beta** | **SE** | **P** | **Beta** | **SE** | **P** | **Z-diff** | **P (adj)** | **Diff** |
| 1 | 55199620 | rs1644513183 | USP24 | -2.728 | 0.400 | 9.46e-12 | -2.159 | 0.491 | 1.09e-5 | -3.854 | 0.684 | 1.77e-8 | 2.013 | 0.618 | No |
| 1 | 1.09E+08 | rs891712338 | CELSR2 | -4.357 | 0.224 | 1.50e-84 | -4.765 | 0.281 | 1.05e-64 | -3.683 | 0.367 | 1.12e-23 | -2.341 | 0.269 | No |
| 2 | 21090558 | rs62122520 | TDRD15 | 2.233 | 0.199 | 3.30e-29 | 2.211 | 0.25 | 8.13e-19 | 2.292 | 0.327 | 2.50e-12 | -0.196 | 1.00 | No |
| 2 | 43839108 | rs2466158484 | ABCG8 | -2.499 | 0.360 | 4.02e-12 | -2.483 | 0.449 | 3.22e-8 | -2.414 | 0.598 | 5.43e-5 | -0.092 | 1.00 | No |
| 5 | 75341886 | rs57479265 | HMGCR | 1.833 | 0.189 | 2.54e-22 | 2.097 | 0.236 | 7.50e-19 | 1.351 | 0.310 | 1.33e-5 | 1.912 | 0.783 | No |
| 6 | 32636278 | rs1018931632 | HLA-DQA1 | 1.779 | 0.312 | 1.21e-8 | 1.488 | 0.391 | 1.43e-4 | 2.221 | 0.513 | 1.51e-5 | -1.134 | 1.00 | No |
| 6 | 1.60E+08 | rs9355746 | IGF2R | -1.202 | 0.207 | 6.55e-9 | -1.181 | 0.259 | 5.29e-6 | -1.296 | 0.341 | 1.46e-4 | 0.267 | 1.00 | No |
| 8 | 1.25E+08 | rs2980886 | TRIB1 | -1.245 | 0.185 | 1.84e-11 | -1.261 | 0.232 | 5.31e-08 | -1.108 | 0.306 | 2.88e-4 | -0.398 | 1.00 | No |
| 9 | 1.33E+08 | rs879055593 | ABO | 1.801 | 0.218 | 1.68e-16 | 1.526 | 0.275 | 3.03e-8 | 2.27 | 0.356 | 1.85e-10 | -1.652 | 1.00 | No |
| 11 | 61834531 | rs1376082791 | FADS2 | -1.446 | 0.212 | 9.48e-12 | -1.367 | 0.266 | 2.83e-7 | -1.616 | 0.349 | 3.59e-6 | 0.568 | 1.00 | No |
| 16 | 56956804 | rs1567455951 | CETP | -1.138 | 0.199 | 1.15e-08 | -1.513 | 0.249 | 1.28e-9 | -0.685 | 0.329 | 3.75e-2 | -2.007 | 0.626 | No |
| 19 | 11080521 | rs2084537196 | LDLR | -5.706 | 0.291 | 9.38e-86 | -5.79 | 0.365 | 1.09e-56 | -5.758 | 0.475 | 8.68e-34 | -0.053 | 1.00 | No |
| 19 | 44908822 | rs3200542 | APOE | -16.872 | 0.344 | 1.0e-300 | -18.218 | 0.428 | 1.0e-300 | -14.455 | 0.569 | 2.81e-142 | -5.283 | **1.78e-6** | **Yes** |
| 20 | 17864040 | rs892381659 | SNX5 | 1.123 | 0.191 | 4.17e-9 | 1.102 | 0.24 | 4.28e-6 | 0.915 | 0.314 | 3.53e-3 | 0.472 | 1.00 | No |

**Table S12. 14 lead genome-wide significant loci identified by positional clumping of combined GWAS results.** Lead SNPs were selected by positional clumping within a 1MB window of combined GWAS genome-wide significant variants

(p < 4.94e-8), retaining the most significant SNP per locus. Effect sizes and p-values are reported for the combined and sex-stratified GWAS. Z-diff denotes the z-score difference statistic testing for sex-differentiated effects, with Bonferroni-corrected

p-values across 14 lead loci (threshold: 3.57e-3).

| **Chr** | **Position** | **Gene** | **Female ꞵ (SE)** | **Male ꞵ (SE)** | **Z-diff** | **P-diff (adj)** | **GxSex ꞵ (SE)** | **GxSex Z** | **P-GxSex(adj)** |
| --- | --- | --- | --- | --- | --- | --- | --- | --- | --- |
| 1 | 55199620 | USP24 | -2.159 (0.491) | -3.854 (0.684) | 2.013 | 0.618 | -0.775 (0.810) | -0.958 | 1.00 |
| 1 | 109274968 | CELSR2 | -4.765 (0.281) | -3.683 (0.367) | -2.341 | 0.269 | 0.758 (0.450) | 1.690 | 1.00 |
| 2 | 21090558 | TDRD15 | 2.211 (0.250) | 2.292 (0.327) | -0.196 | 1.00 | 0.039 (0.415) | 0.094 | 1.00 |
| 2 | 43839108 | ABCG8 | -2.483 (0.449) | -2.414 (0.598) | -0.092 | 1.00 | -0.169 (0.731) | -0.232 | 1.00 |
| 5 | 75341886 | HMGCR | 2.097 (0.236) | 1.351 (0.310) | 1.912 | 0.783 | -0.619 (0.390) | -1.590 | 1.00 |
| 6 | 32636278 | HLA-DQA1 | 1.488 (0.391) | 2.221 (0.513) | -1.134 | 1.00 | 0.446 (0.649) | 0.687 | 1.00 |
| 6 | 160103304 | IGF2R | -1.181 (0.259) | -1.296 (0.341) | 0.267 | 1.00 | 0.057 (0.424) | 0.135 | 1.00 |
| 8 | 125467846 | TRIB1 | -1.261 (0.232) | -1.108 (0.306) | -0.398 | 1.00 | 0.227 (0.380) | 0.598 | 1.00 |
| 9 | 133271182 | ABO | 1.526 (0.275) | 2.270 (0.356) | -1.652 | 1.00 | 0.405 (0.450) | 0.900 | 1.00 |
| 11 | 61834531 | FADS2 | -1.367 (0.266) | -1.616 (0.349) | 0.568 | 1.00 | -0.293 (0.434) | -0.674 | 1.00 |
| 16 | 56956804 | CETP | -1.513 (0.249) | -0.685 (0.329) | -2.007 | 0.626 | 0.719 (0.411) | 1.750 | 1.00 |
| 19 | 11080521 | LDLR | -5.790 (0.365) | -5.758 (0.475) | -0.053 | 1.00 | -0.133 (0.575) | -0.231 | 1.00 |
| 19 | 44908822 | APOE | -18.218 (0.428) | -14.455 (0.569) | -5.283 | 1.78e-6 | 3.541 (0.657) | 5.387 | 1.01e-6 |
| 20 | 17864040 | SNX5 | 1.102 (0.240) | 0.915 (0.314) | 0.472 | 1.00 | -0.175 (0.394) | -0.444 | 1.00 |

**Table S13: Concordance of GxSex interaction estimates with the sex-stratified z-score difference test of statin-adjusted LDL-C.** Female and male ꞵ (SE) are the effects on statin-adjusted LDL-C (mg/dL) from the sex-stratified GWAS. Z-diff denotes the z-score difference statistic testing for sex-differentiated effects. GxSex ꞵ (SE) and GxSex Z are for the SNP-by-sex term from the GxSex model in the sex-pooled sample with females as the reference (male-minus-female), so their signs are opposite to Z-Diff. P-diff and P-GxSex are Bonferroni-corrected p-values across 14 lead loci (threshold: 3.57e-3).

| **Statin Use Status** | **LDL-C Phenotype** | **Female Estimate** | **Female SE** | **Male Estimate** | **Male SE** | **Z-Diff** | **P-Value** |
| --- | --- | --- | --- | --- | --- | --- | --- |
| **Heritability (V(G)/Vp)** | | | | | | | |
| Untreated | Unadjusted | 0.041 | 0.011 | 0.028 | 0.016 | 0.695 | 0.487 |
| Statin-treated | Adjusted | 0.037 | 0.036 | 0.042 | 0.033 | -0.089 | 0.929 |
| Statin-treated | Unadjusted | 0.037 | 0.036 | 0.042 | 0.033 | -0.089 | 0.929 |
| **Additive Genetic Variance (V(G))** | | | | | | | |
| Untreated | Unadjusted | 42.100 | 10.900 | 29.900 | 17.600 | 0.5870 | 0.557 |
| Statin-treated | Adjusted | 73.300 | 70.700 | 68.900 | 55.100 | 0.0484 | 0.961 |
| Statin-treated | Unadjusted | 35.900 | 34.700 | 33.800 | 27.000 | 0.0484 | 0.961 |
| **Environmental Variance (V(e))** | | | | | | | |
| Untreated | Unadjusted | 985.000 | 13.900 | 1057.000 | 21.700 | -2.790 | 0.005 |
| Statin-treated | Adjusted | 1884.000 | 78.100 | 1580.000 | 61.200 | 3.070 | 0.002 |
| Statin-treated | Unadjusted | 923.000 | 38.300 | 774.000 | 30.000 | 3.070 | 0.002 |
| **Phenotypic Variance (Vp)** | | | | | | | |
| Untreated | Unadjusted | 1028.000 | 9.630 | 1087.000 | 13.600 | -3.590 | 3.33e-4 |
| Statin-treated | Adjusted | 1957.000 | 36.400 | 1649.000 | 29.400 | 6.590 | 4.50e-11 |
| Statin-treated | Unadjusted | 959.000 | 17.800 | 808.000 | 14.400 | 6.590 | 4.50e-11 |

**Table S14: Heritability and variance estimates of LDL-C for women and men.** Heritability and variance component estimates and standard errors for women and men as output by GCTA GREML. Individuals were stratified by sex then by statin usage status. Heritability of unadjusted LDL-C was estimated for all groups, and additionally statin-adjusted LDL-C for statin-treated individuals. Z-Diff and P-Value denote the z-score difference and respective p-value between women and men within each stratum.

| **Statin Use Status** | **LDL-C Phenotype** | **Female Estimate** | **Female SE** | **Male Estimate** | **Male SE** | **Z-Diff** | **P-Value** |
| --- | --- | --- | --- | --- | --- | --- | --- |
| **Heritability (V(G)/Vp)** | | | | | | | |
| Untreated | Unadjusted | 0.064 | 0.011 | 0.058 | 0.018 | 0.292 | 0.770 |
| Statin-treated | Adjusted | 0.068 | 0.053 | 0.020 | 0.043 | 0.704 | 0.482 |
| Statin-treated | Unadjusted | 0.068 | 0.053 | 0.020 | 0.043 | 0.704 | 0.482 |
| **Additive Genetic Variance (V(G))** | | | | | | | |
| Untreated | Unadjusted | 75.200 | 13.200 | 76.300 | 23.500 | -0.041 | 0.968 |
| Statin-treated | Adjusted | 219.000 | 171.000 | 61.600 | 134.000 | 0.727 | 0.467 |
| Statin-treated | Unadjusted | 108.000 | 83.600 | 30.200 | 65.700 | 0.727 | 0.467 |
| **Environmental Variance (V(e))** | | | | | | | |
| Untreated | Unadjusted | 1095.000 | 16.100 | 1237.000 | 27.400 | -4.450 | 8.430e-6 |
| Statin-treated | Adjusted | 3021.000 | 181.000 | 3046.000 | 148.000 | -0.104 | 0.917 |
| Statin-treated | Unadjusted | 1480.000 | 88.800 | 1492.000 | 72.300 | -0.104 | 0.917 |
| **Phenotypic Variance (Vp)** | | | | | | | |
| Untreated | Unadjusted | 1170.000 | 11.100 | 1313.000 | 16.400 | -7.210 | 5.49e-13 |
| Statin-treated | Adjusted | 3241.000 | 72.700 | 3107.000 | 64.700 | 1.370 | 1.700e-1 |
| Statin-treated | Unadjusted | 1588.000 | 35.600 | 1522.000 | 31.700 | 1.370 | 1.700e-1 |

**Table S15: Heritability and variance estimates of maximum LDL-C for women and men.** Heritability and variance component estimates and standard errors for women and men as output by GCTA GREML for maxim LDL-C. Individuals were stratified by sex then by statin usage status. Heritability of unadjusted LDL-C was estimated for all groups, and additionally statin-adjusted LDL-C for statin-treated individuals. Z-Diff and P-Value denote the z-score difference and respective p-value between women and men within each stratum.
